# Altered skin microbiome, inflammation, and JAK/STAT signaling in Southeast Asian ichthyosis patients

**DOI:** 10.1101/2022.08.02.22277646

**Authors:** Minh Ho, Huynh-Nga Nguyen, Minh Van Hoang, Tien Thuy Thi Bui, Bao-Quoc Vu, Truc Huong Thi Dinh, Vo Hoa Thi My, Diana Blaydon, Sherif A. Eldirany, Christopher G. Bunick, Chi-Bao Bui

## Abstract

Cutaneous ichthyosis (CI) is a collective group of monogenetic disorders of cornification demonstrating epidermal scaling, fissuring, chronic skin inflammation, and increased susceptibility to infection. In healthy individuals the skin microbiome limits growth of pathogenic organisms; however, the microbiome signature in CI is poorly characterized. To rectify this, we investigated the microbiome signature across 7 subtypes of CI in 43 individuals of Southeast Asian ethnicity, of which exome sequencing revealed 20 novel and 31 recurrent pathogenic variants. Microbiome meta-analysis revealed distinct microbial populations, reduced commensal microbiota, and higher colonization by pathogenic species. This correlated with increased production of inflammatory cytokines, including Th17 and JAK/STAT signaling, in peripheral blood mononuclear cells. Moreover, we identified microbiota and inflammation alterations in wounds of CI patients responsible for impaired wound healing. Together, this research enhances our understanding of the microbiological, immunological, and molecular properties of CI patients and provides critical information for improving therapeutic management.

## Introduction

Ichthyosis originates from the Greek word “*ichthys*”, meaning “fish”, to describe a fish-scale-like skin phenotype in humans. Human ichthyoses, of which there are ∼20 types, are predominantly inherited disorders divided into non-syndromic or syndromic forms; they are characterized by malfunction of keratinocyte proteins, lipid biosynthesis, cell adhesion, and DNA repair [1]. Cutaneous ichthyosis (CI) often displays constant scaling of the skin, but some forms are associated with a severe, often fatal, neonatal outcome due to infection while having an immature immune system. The severity of CI ranges from dry skin (xerosis cutis) in ichthyosis vulgaris (IV) to life-threatening Harlequin ichthyosis (HI). The compromised epidermal barrier in CI patients (e.g., chronic scaling and fissuring) may facilitate access of skin bacteria and/or fungi into sub-corneal layers via scratch-related lesions leading to recurrent/chronic skin wounds, inflammation, and high risk of cutaneous and systemic infections.

Human skin is a crucial physical barrier against infection, water loss, and body temperature dysregulation. It also hosts a diverse, symbiotic microbiome. The human skin microbiome predominantly belongs to the phyla *Actinobacteria*, *Firmicutes*, *Proteobacteria*, and *Bacteroidetes*[2], and they work cooperatively with biproducts of the stratum corneum, like neutral/polar lipids and glycosphingolipids [3], to prevent cutaneous domination by infectious pathogenic microbes[4, 5]. However, the balance or homeostasis can be disrupted by mutations/alterations in the human epidermis [6] or skin barrier processing proteins[7], increasing the potential for infection and phenotypic diseases like atopic dermatitis (AD) or a type of CI. Cohort studies of AD demonstrated that AD severity correlated with the dominance of *Staphylococcus aureus* over *S. epidermidis* [8], as well as reduction of cutaneous microbiome diversity [9]. This illustrates how the resident microbiome’s composition can influence skin disease activity. Despite a number of studies evaluating AD [6, 8, 10–13] and psoriasis [14–16] skin microbiome compositions with proposed therapeutic treatments [17, 18], the cutaneous microbiome across the various CI types has not been well-characterized.

Recently, immunology profiling of ichthyosis demonstrated that Th17 signaling is elevated especially with the involvement of IL-17 for Congenital Ichthyosiform Erythroderma (CIE), Lamellar Ichthyosis (LI), and Epidermolytic Ichthyosis (EI) [19–21]. Guttman-Yassky and colleagues showed the immune profile and lipid metabolism of ichthyosis (CIE, LI, HI) is more similar to psoriasis than AD, indicating the potential for psoriasis biologics as viable therapies for ichthyosis patients. Interestingly, the severity of AD correlated more with cytokine expression rather than with the exact type of gene mutation [22]. This data exposes an important knowledge gap – what is the connection between the microbiome, immune profile, and disease pathogenesis in CI? Currently the clinical evaluation, diagnosis, and management of CI utilizes serological testing, histology, bacterial cultures, and incidence of viral and fungal infections; however, the lack of extensive characterization of the skin microbiome associated with CI disorders limits accurate diagnosis and effective treatment.

The work reported here advances CI biology in several ways: It provides a comprehensive characterization of the microbiome signature across 7 groups of CI patients (IV, Trichothiodystrophy (TTD), EI, LI, Arthrogryposis renal dysfunction cholestasis (ARC), Sjögren-Larsson Syndrome (SLS), and HI); it correlates genotype with clinical phenotype for 31 pathogenic/likely pathogenic and 20 novel variants (previously unreported mutations in ichthyosis patients); and it establishes an immunophenotype and microbiome signature for CI patients during the wound healing process. First, a cohort of 36 CI patients and 7 healthy controls were followed long-term (median time 4.3 years) over which clinical phenotype was documented and genotype analyzed by whole exome sequencing. Second, microbiome sequencing was performed for dry, moist, and sebaceous skin sites. Third, using principal coordinate analysis on the microbiome signature the cohort was separated into 5 dysbiosis clusters and further characterized for microbiota shifts during wound healing, wound healing response to antibiotic therapy, and immunophenotyping in unwounded and wounded states. Together this research enhances our understanding of the microbiological, immunological, and molecular properties of CI patients, with emphasis on ethnic (Asian) skin. Such emphasis is important since racial differences in skin barrier function are reported, especially with Asian skin having higher rates of transepidermal water loss and skin reactivity compared to black or Caucasian skin [23].

## Results

### Characterization of clinical and genetic variants in CI patients

A study cohort of 43 individuals of Southeast Asian ethnicity (36 CI patients plus 7 age-matched healthy subjects) were assessed clinically and further analyzed by microbiome sequencing, whole exome sequencing (WES), and immunophenotyping (Fig. 1A). First, we examined the clinical patterns of each disorder and compared them to related genetic mutations as defined and categorized by expert dermatologists, microbiologists, and geneticists in Vietnam. We categorized the CI patients (n = 36) into having one of 7 CI disorders: IV (n=15), HI (n=8), LI (n=3), EI (n=4), TTD (n=3), ARC (n=1), and SLS (n=2) and compared them alongside healthy age-matched subjects (n = 7). The median duration of follow-up was 4.3 years with patient age (average 10.2 years old) ranging from neonatal (birth) to adults (40 years). Clinical phenotypes of some of the CI patients are shown (Fig. 1B).

**Figure 1.**
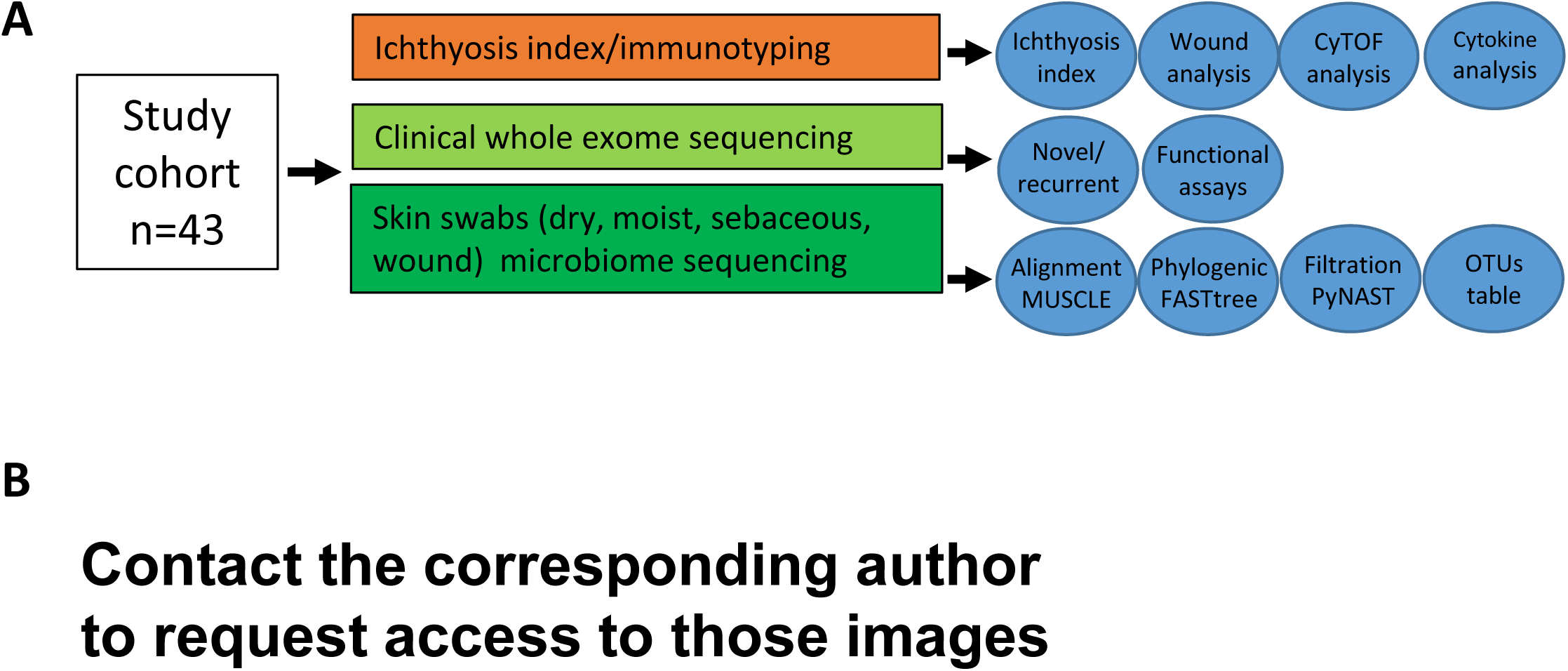
Study design and clinical phenotypes found in Southeast Asian individuals with cutaneous ichthyosis (CI). A. Schematic overview of study design and methods used. B. Clinical photographs of representative individuals with harlequin ichthyosis (HI1, HI5, HI7, HI8), lamellar ichthyosis (LI1), epidermolytic ichthyosis (EI1), trichothiodystrophy/XPD (TTD1), ichthyosis vulgaris (IV1), arthrogryposis, renal dysfunction, and cholestasis (ARC1) syndrome. * Contact the corresponding author to request access to those images

To characterize the frequency of specific clinical manifestations in CI, our cohort was evaluated for ten findings: hyperkeratosis, itch, microbial infection, erythroderma, respiratory difficulties, sepsis, collodion membrane at birth, loss of heat, blistering, and cardiac abnormalities. Hyperkeratosis and itch were the most common clinical manifestations of CI (100% and 97.2%, respectively), followed by microbial infection (94.4%) and erythroderma (72.2%) (Table 1). 52.78% of CI patients demonstrated respiratory comorbidities and/or sepsis. There were 6 lethal cases due to infant sepsis (4 HI, 1 LI, and 1 EI). Ten other cases of childhood sepsis were evident in HI (3 out of 8) or in IV (7 out of 15), but were treatable with antibiotics (Tables S1 and S2). Most HI and LI cases also presented with neonatal-onset chronic diarrhea, episodes of fever, recurrent pyodermas, oral candidiasis, and otitis externa. Aside from sepsis, one TTD patient developed necrotizing fasciitis of the nose following *Burkholderia pseudomallei* infection.

**Table 1.** Clinical phenotypes of cutaneous ichthyosis (CI) cohort.

| Clinical manifestations | Patients (# of CI subtype patients with clinical finding) (n=36) | Frequency |
| --- | --- | --- |
| Hyperkeratosis (thick skin) | IV(15), HI(8), LI(3), EI(4), TTD(3), ARC(1), SLS (2) | 100% |
| Itch/Pruritus | IV(15), HI(8), LI(3), EI(4), TTD(3), ARC(1), SLS (1) | 97.22% |
| Skin microbial infection* (n=18) | IV(9), HI(2), EI(1), TTD(2), SLS (1) | 83.33% |
| Erythroderma | IV(10), HI(8), LI(1), EI(4), TTD(2) SLS (1) | 72.22% |
| Respiratory comorbidities | IV(10), HI(3), EI(2), TTD(3), SLS (1) | 52.78% |
| Sepsis | IV(7), HI (6), LI(2), EI(1), TTD(3) | 52.78% |
| Collodion membrane at birth | IV(1), HI(8), LI(3) | 33.33% |
| Cardiomyopathy comorbidities | IV(4), HI (2), EI(2), TTD(3) | 30.56% |
| Blistering | HI(8), EI(2) | 27.78% |
| Loss of heat | HI(3), LI(1), EI(2) | 16.67% |

The total number of CI patients (n=36) in our cohort were categorized as: ichthyosis vulgaris (IV, n=15), harlequin ichthyosis (HI, n=8), lamellar ichthyosis (LI, n=3), epidermolytic ichthyosis (EI, n=4), trichothiodystrophy (TTD, n=3), arthrogyrosis, renal dysfunction, and cholestasis (ARC) syndrome (n=1), and Sjögren–Larsson syndrome (SLS, n=2). \*Only n=18 patients involved in testing for this manifestation; data of microbial infection and loss of heat shown in Tables S2 and S3, respectively.

Next, we performed WES and genetic linkage analysis in the 36 CI individuals. 31 pathogenic/likely pathogenic mutations (based on American College Medical Genetics) and 20 novel variants were identified (Table 2). Four novel variants were found in more than one CI patient in our cohort. We identified compound heterozygous and homozygous segregated mutations linked to autosomal recessive loss-of-function of *ABCA12, TGM1, ALDH3A2*, and *ERCC2.* The genetic mutations are mapped onto their corresponding protein domain (Fig. 2) and where possible, we analyzed the protein structure alterations caused by the CI-related missense mutations (Figs. S1-S4).

**Figure 2.**
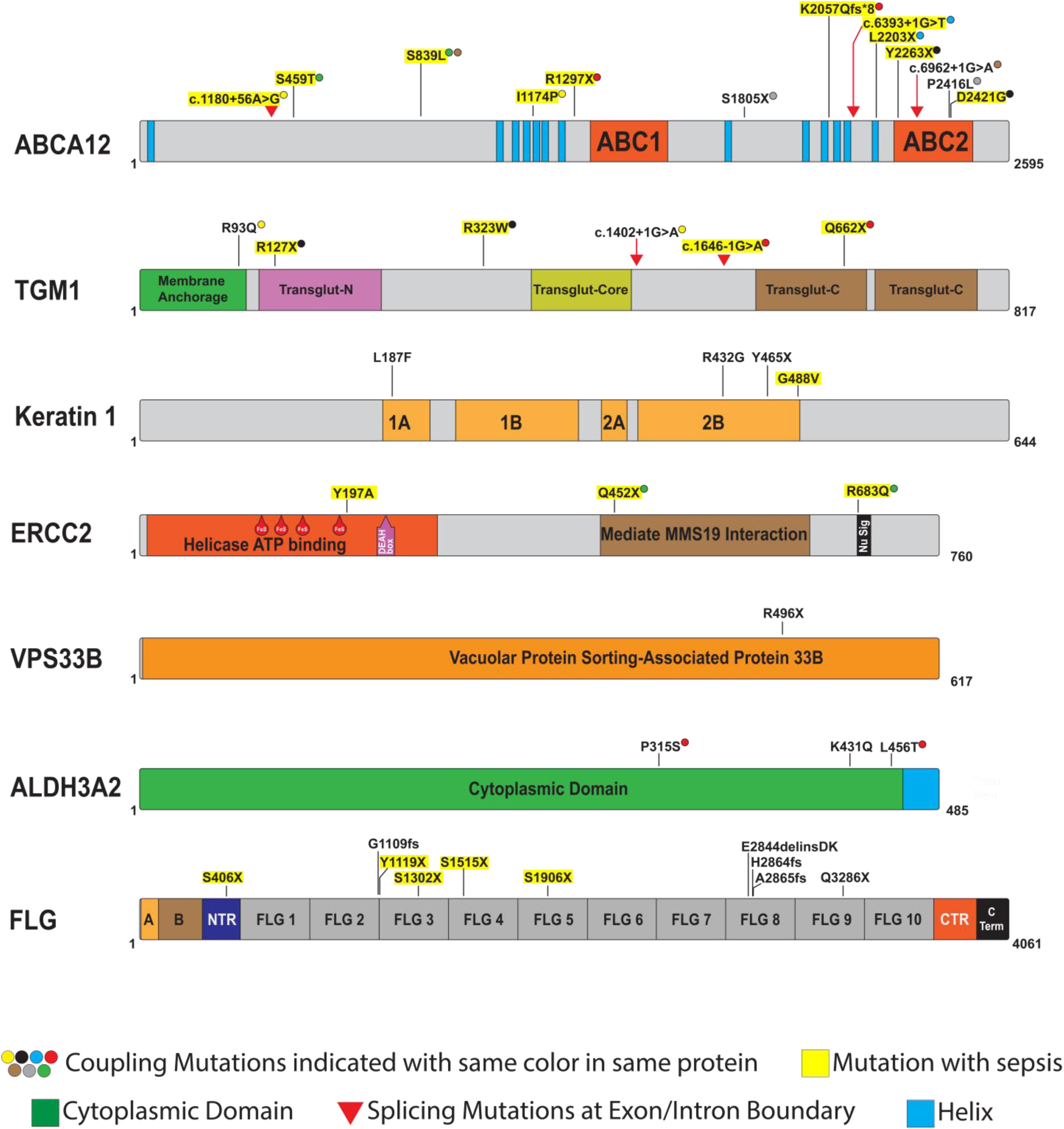
Pathogenic variants identified in cohort of Southeast Asian CI patients. Schematic representation of mutated gene domains with annotated missense, nonsense, splicing, and frameshift mutations identified in affected individuals with *ABCA12, TGM1, KRT1, FLG, ERCC2, VSP33B*, and *ALDH3A2* mutations. “Coupling mutations” refers to variants identified together in the same patient. Variants highlighted with yellow experienced sepsis: we note that 70% of the ABCA12 variants associated with sepsis in our cohort were adjacent to the ABC1 or ABC2 domains; KRT1 G488V variant associated with sepsis occurred at the C-terminus in the highly conserved TYR*LLEGE motif known to be critical for intermolecular and higher order filament interactions (see Fig. S2); IV patients who experienced sepsis had either frameshift or nonsense mutations in the N-terminal half of *FLG* whereas those having mutations more C-terminal did not experience sepsis.

**Table 2.**
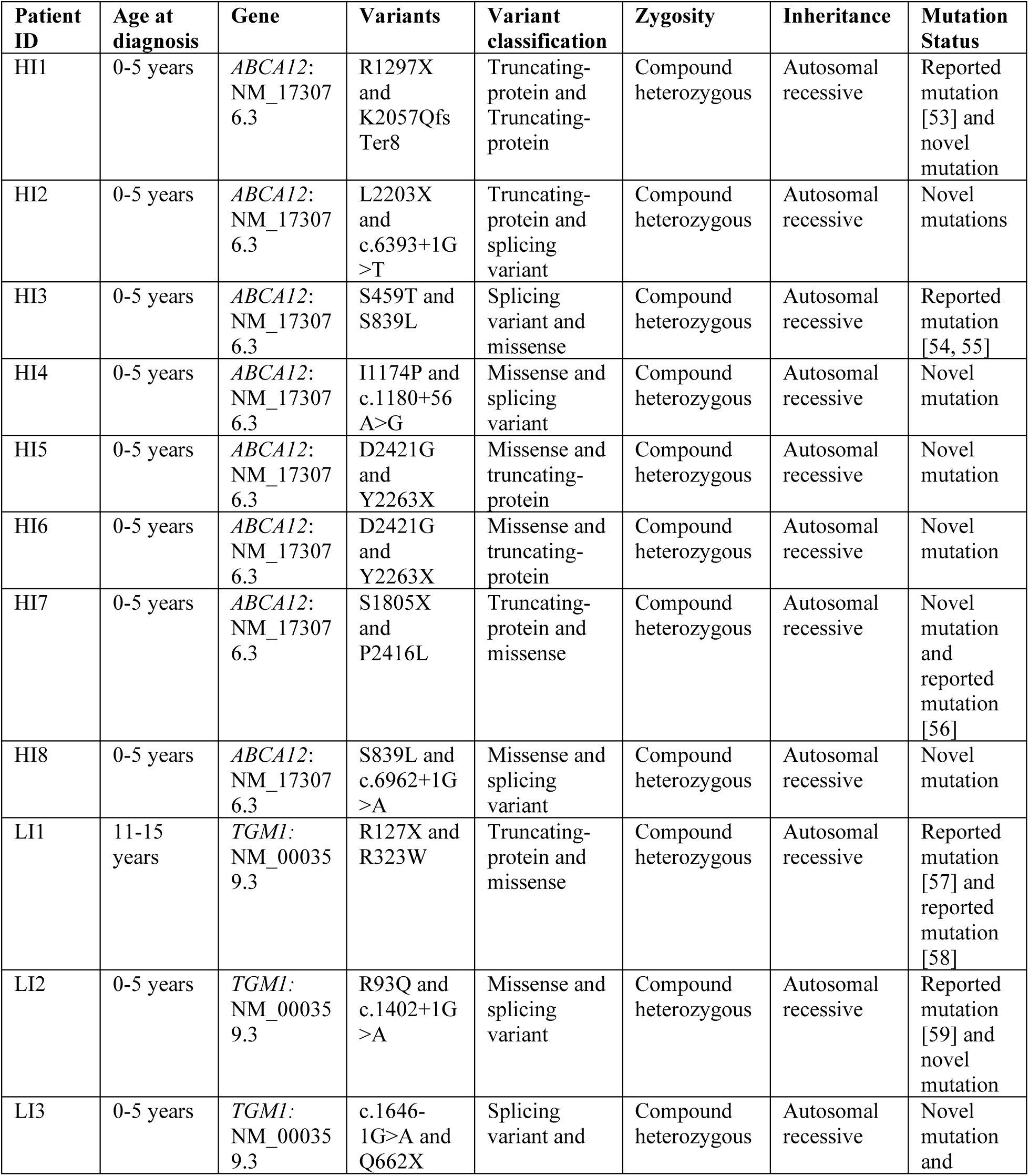
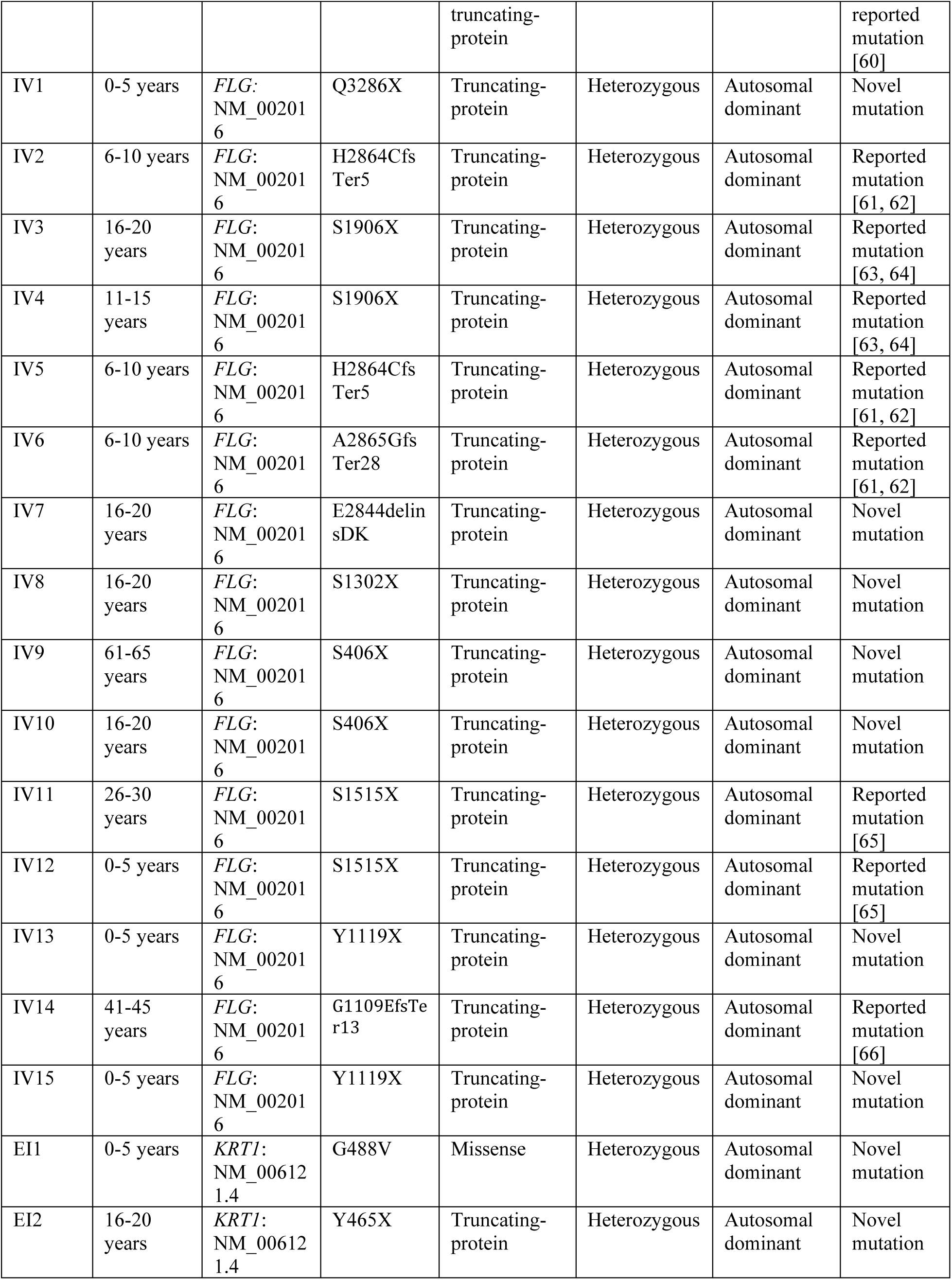
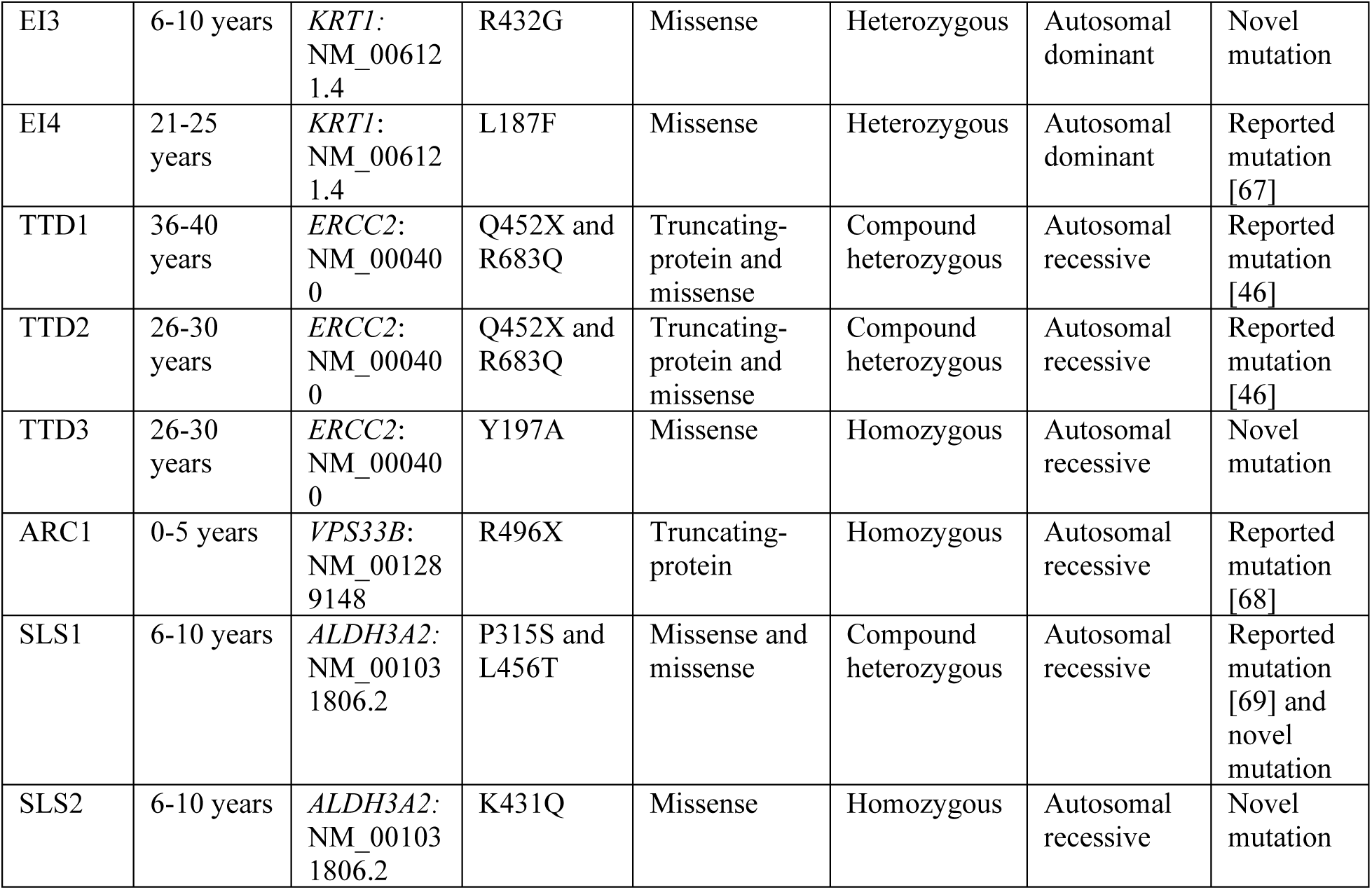
Whole exome sequencing (WES) analysis of 36 CI patients. For each CI patient, age at diagnosis, gene name and sequence identifier, amino acid variant, variant classification, mode of inheritance, and novelty of mutation are presented.

### CI patient microbiome signatures

To better understand the risk of infections in CI, we comprehensively examined the CI patient microbiome (n=36) by taking superficial swabs of sebaceous skin (face), dry skin (leg) and moist skin (arm) and compared the results to age-matched healthy subjects (n=7; Fig. 1A). We focused on the four dominant phyla - Actinobacteria, Firmicutes, Proteobacteria, and Bacteroidetes. Actinobacteria and Firmicutes both show decreased average relative abundance of colony forming units (cfu) in CI dry, moist, and sebaceous skin compared to the respective locations in healthy controls (Fig. 3A). Proteobacteria showed decreased relative abundance of cfu in dry CI skin compared to healthy controls, but increased abundance in moist and sebaceous skin. Bacteroidetes appeared stable to slightly increased in relative abundance of cfu across all skin locations in CI patients compared to healthy controls.

**Figure 3.**
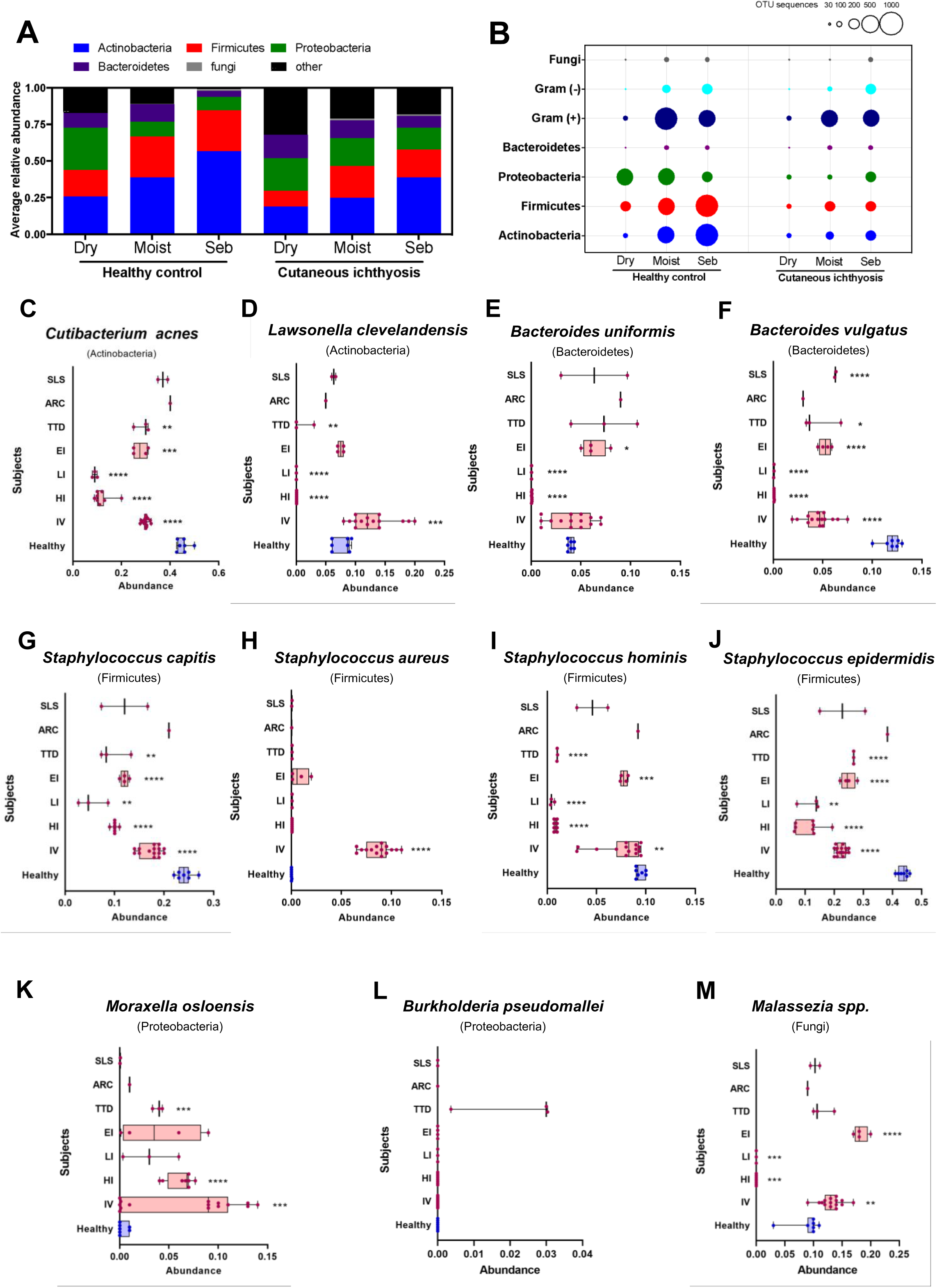
Microbiome profile in CI patients. A. Bacterial community composition at phylum level (Actinobacteria, Firmicutes, Proteobacteria, Bacteroidetes, fungi, and other) represented as average relative abundance from healthy (left) and CI patients (right) and three different skin sites: dry, moist, and sebaceous. B. Relative abundance of operational taxonomic units (OTUs) in healthy (left) and CI patients (right) from three different skin sites (dry, moist, and sebaceous) by White’s non-parametric t-test. Circle sizes are proportional to the number of sequences within for the phyla Actinobacteria, Firmicutes, Proteobacteria, Bacteroidetes, fungi, and gram positive/gram negative bacteria. C-M. Abundance of specific bacterial species for each CI clinical type: C, *Cutibacterium acnes*; D, *Lawsonella clevelandensis*; E, *Bacteroides uniformis*; F, *Bacteroides vulgatus*; G, *Staphylococcus capitis*; H, *Staphylococcus aureus*; I, *Staphylococcus hominis*; J, *Staphylococcus epidermidis;* K, *Moraxella osloensis*; L, *Burkholderia pseudomallei*; M, *Malassezia* species. Results are expressed in dot plot with median and interquartile range with statistical significance as *\*p* ≤ 0.05, *\*\*p* ≤ 0.01, *\*\*\*p* ≤ 0.001, *\*\*\*\*p* ≤ 0.0001.

Next, we compared the operational taxonomic units (OTU) sequence reads between the CI patients (n=36) and healthy controls (n=7) for dry, moist, and sebaceous skin locations (Fig. 3B). We observed reduced OTU counts of Actinobacteria, Firmicutes, and Proteobacteria across dry, moist and sebaceous skin, but no significant difference in Bacteroidetes OTU counts (Fig. 3B). Analysis of the OTU sequences at the species level identified a significant reduction in species within the Actinobacteria, Firmicutes, and Bacteroidetes phyla for certain CI types (Figs. 3C-L). For Actinobacteria, CI patients generally had reduced levels of *Cutibacterium acnes* and *Lawsonella clevelandensis* compared to healthy controls, with the reduction in *C. acnes* being statistically significant for IV, HI, LI, EI, and TTD types (Fig. 3C). *L. clevelandensis* demonstrated a mixed pattern, with significant elevations in IV patients but significant decreases in LI, HI, and TTD patients (Fig. 3D). For the Bacteroidetes phylum, *Bacteroides uniformis* had significant reductions in LI and HI patients, and *B. vulgatus* had significant reductions in IV, HI, LI, EI, TTD, and SLS patients (Fig. 3E, F). Similarly, *Staphylococcus capitis, S. hominis,* and *S. epidermidis* of the Firmicutes phylum showed significantly reduced levels in IV, HI, LI, EI, and TTD compared to healthy controls (Fig. 3G-J).

In contrast to the reductions in *C. acnes*, *S. capitis*, *B. uniformis*, and *B. vulgatus*, *S. aureus* of the Firmicutes phylum showed statistically significant elevations in IV patients versus healthy controls, and statistically non-significant elevations in EI patients (Fig. 3H). Soil-dwelling and zoonotic disease-associated bacteria from the Proteobacteria and Actinobacteria phyla were also identified in certain CI patients. *Burkholderia pseudomallei* was exclusively increased in TTD patients (Fig. 3L), whereas *Moraxella osloensis* was increased in multiple CI types, most significantly TTD, HI, and IV (Fig. 3K). Lastly, LI and HI were unique among the CI types in demonstrating a marked reduction in *Malassezia* species, in contrast to EI and IV patients who experienced higher *Malassezia* abundance (Fig. 3M).

### Principal co-ordinate analysis identifies CI microbiome subgroups

To investigate differences in pathogenic and commensal bacteria in healthy and CI patients across skin locations, the superficial swab OTU sequence data was subjected to principal co-ordinate analysis (PCA). We identified dysbiosis clusters by collecting high-ratio non-pathogenic (principal component 1, PC1, x-axis) and high ratio pathogenic (principal component 2, PC2, y-axis) microbiomes observed in CI patients (Fig. 4A). Patients segregated into five distinct clusters following analysis of dry, moist, and sebaceous skin samples: cluster P1–healthy control subjects were high in PC1 and low in PC2; cluster P2–IV patients were high in PC2 and low in PC1 (*FLG* variants); cluster P3–EI and TTD patients were low in PC1 and variably elevated in PC2 in moist and sebaceous skin (*KRT1* and *ERCC2* variants, respectively); cluster P4–LI and HI patients were consistently lowest in both PC1 and PC2 across all skin sites (*TGM1* and *ABCA12* variants, respectively); and cluster P5–ARC and SLS patients were low in PC1 and variably elevated in PC2 in sebaceous skin only (*VPS33B* and *ALDH3A2* variants, respectively) (Fig. 4B).

**Figure 4.**
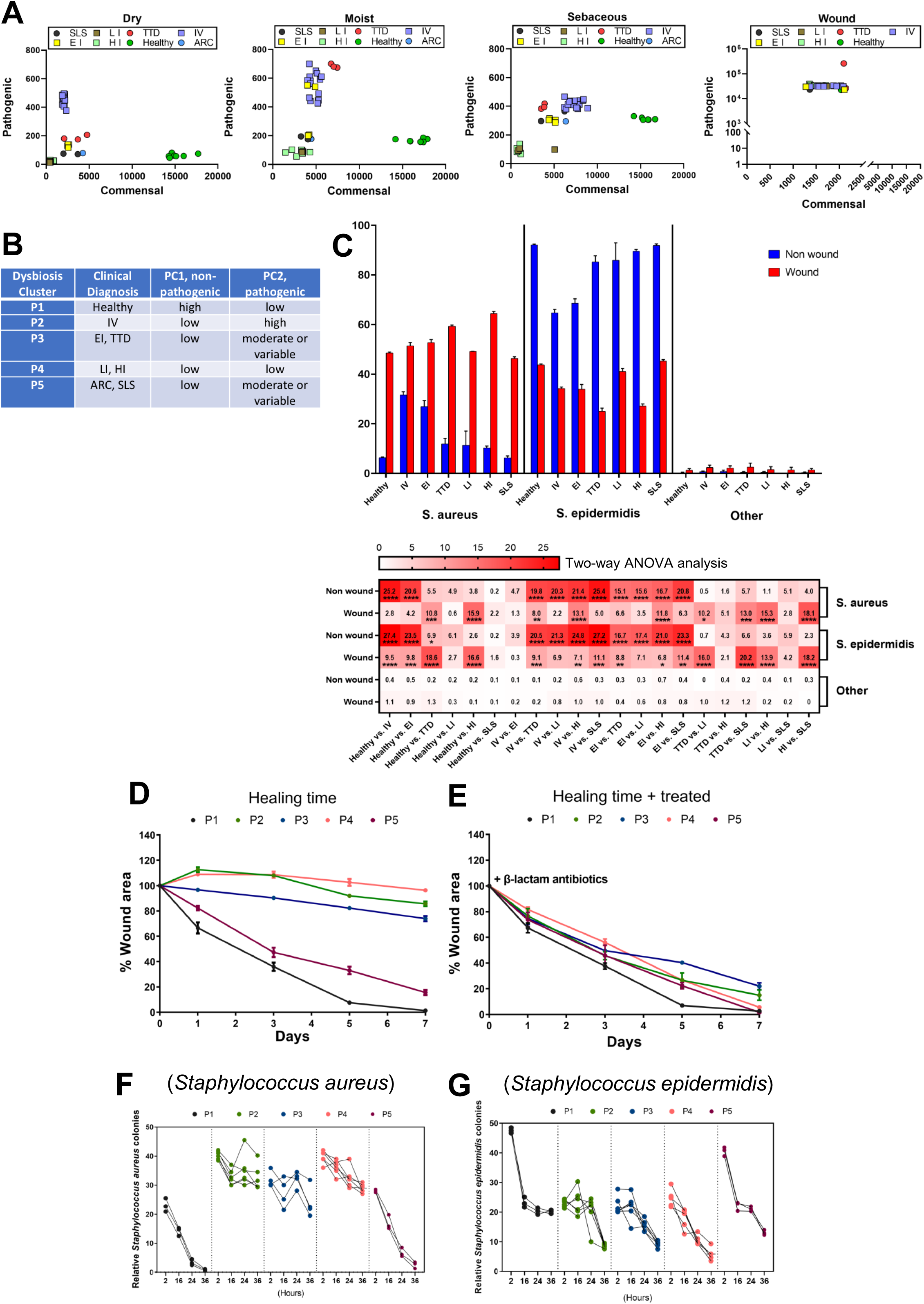
Dysbiosis clustering of CI patients and the pathogenic microbiome in wounded skin of CI patients. A. PCoA plots showing beta diversity metrics in dry, moist, sebaceous and wounded CI skin, colored according to CI type. B. CI microbiome dysbiosis clusters P1-P5 as determined by principal co-ordinate analysis in panel A (PC1/commensal, PC2/pathogenic). C. (top) Dynamic changes in abundance of *S. aureus and S. epidermis* communities in non-wounded (blue) and wounded (red) skin from representative CI patients IV, EI, TTD, LI, HI, SLS compared with healthy controls. (bottom) Heatmap diagram showing each CI group compared to healthy controls, and each group compared each other, using two-way ANOVA analysis. Results of each bar are expressed as mean ± SEM standard error. *\*p* ≤ 0.05, *\*\*p* ≤ 0.01, *\*\*\*p* ≤ 0.001, *\*\*\*\*p* ≤ 0.0001. D, E. After wounding CI patients and healthy controls with 5 mm punch biopsy, healing time, measured as the change in percent wound area over 7 days, was delayed for the 4 CI dysbiosis clusters P2-P5 compared to healthy P1 controls in the absence of antibiotics (D), but in the presence of β-lactam antibiotics healing time improved to levels comparable to healthy controls in (E). F, G. Measurement of relative colonies of *Staphylococcus aureus* (F) or *Staphylococcus epidermidis* (G) across the 5 dysbiosis clusters in wounded skin over 36 hours.

### Wound infection impairs healing of CI skin

To investigate how the microbiota changes in CI patients when their skin is wounded, we introduced 5 mm punch biopsy wounds in a CI wound-cohort (n=32; the ARC patient from P5 did not participate) and matched healthy controls (n=5) and evaluated them for microbial infection, wound healing time, and inflammation. Using PCA on collected OTU sequences, we demonstrated that wounds in CI skin are characterized by a major shift in the dysbiosis clusters of CI patients to a very high PC2 and low PC1 state (Fig. 4A, far right panel). This same shift occurred for the healthy controls.

Next, we delineated the relative abundance of microbial species in CI wounds. In non-wound skin, *S. epidermidis* abundance was high for healthy and CI skin, with EI and IV patients having the least abundant *S. epidermidis* (Fig. 4C blue). In non-wound skin *S. aureus* abundance was much lower than *S. epidermidis*, though higher among CI types than healthy controls. In contrast, wounded skin demonstrated a large shift towards more abundant *S. aureus* (mean = 48.5% total relative abundance) for healthy and CI patients, with concomitant reduction in *S*. *epidermidis* (mean = 43.76% total relative abundance) (Fig. 4C, red), as supported by two-way ANOVA analysis. *Streptococcus pyogenes* (mean = 3.3% total relative abundance) and low proportions of the zoonotic pathogen *Streptococcus suis* (0.3% relative abundance) were also observed more in wounded skin compared to healthy control skin (Fig. S5A).

Some Gram-negative, facultative anaerobes such as *Pseudomonas aeruginosa* and *Campylobacter* demonstrated different relative abundance among dysbiosis clusters. For example, *P. aeruginosa* (2.15% relative abundance) showed the greatest elevated abundance in P2 (IV) and P3 (EI/TTD), a smaller elevation in P5 (SLS), but was not detected in P4 (LI/HI), compared to healthy controls (Fig. S5A). *Campylobacter* (0.73% relative abundance) similarly showed elevated abundance in P2, P3, and the LI patients in P4. *Candida albicans* (0.82% relative abundance) was similar in abundance across healthy controls and IV patients (P2), showed increased abundance in EI patients (P3) and SLS patients (P5), but showed decreased abundance in P4 and among TTD patients (P3). Methicillin-resistant *S. aureus* (MRSA) was most abundant in HI patients (P4) (Figs. S5A and S6A), and also observed in IV (P2). Using real-time PCR, we observed elevated HPV virus (1.45% relative abundance) in TTD (P3) and LI (P4) patients (Fig. S6B).

To further evaluate whether the abundance of pathogenic microbes in CI patients (P2, P3, P4, and P5) impairs wound healing time versus healthy control subjects (P1), we wounded by 5 mm punch biopsy CI and control patients and observed their wound healing times in the absence or presence of oral 𝛃-lactam antibiotics (carbapenems and cephalosporins) (Figs. 4D, E). In the absence of antibiotics, the time required for wound healing was significantly delayed in P2, P3, and P4 CI patients compared to healthy controls (P1), with a smaller degree of wound healing delay in P5. In contrast, administration of antibiotics led to nearly complete wound healing in all 4 CI dysbiosis clusters (P2-P5), with little change in the trajectory of healing time by healthy controls, indicating that the altered CI microbiome impairs wound healing.

To further characterize the response of wounded skin in CI patients, we evaluated expression levels of the anti-microbial peptide 𝛃-Defensin 2 (HBD-2) both in healthy subjects (P1) and CI patients (P2-P5) (Fig. S5B). We observed elevations of HBD-2 mRNA in wounded skin of CI patients from dysbiosis clusters P2, P3, and P4 compared to wounded skin of healthy controls (P1). CI patients in cluster P5, however, had similar levels of HBD-2 mRNA in wounded skin as the healthy controls. *S. aureus* colony counts (CFU) were significantly reduced in the wounded skin of healthy subjects (P1) and P5 CI patients after approximately 36 hours of healing time (Fig. 4F). *S. aureus* colony counts remained high in P2-P4 CI patients after 36h. *S. epidermidis* colony counts among healthy subjects (P1) and CI patients (P2-P5) were similarly reduced after a wound healing time of 24-36h (Fig. 4G), although healthy subjects started and ended with higher overall *S. epidermidis* colony counts. *P. aeruginosa* colony counts were highly increased at 2h and reduced back at 16h in P3 and P4 CI patient wounds, and to lesser extend in P5, from 2 to 16 hours healing time (Fig. S5C)

In summary, CI patients showed: (i) distinct microbiota signatures with high dysbiosis, (ii) pathogenic bacterial levels linked to IV (P2) and EI and TTD (P3) patients, and (iii) generally low levels of all microbes present in HI and LI patients (P4). Viral susceptibility, such as to HPV or HBV/HCV, is particularly linked with CI patients (Table S2). Breached wounds are a key determinant of all infectious microbe levels, mainly in the ratio of *S. aureus* to *S. epidermis* (Fig. 4C and Table S1) and may drive the sepsis seen in CI patients. Therapies for CI skin can correct the ratio of *S. aureus* to *S. epidermis*, as we observed for TTD patients receiving phototherapy and IV patients receiving topical ammonium bituminosulfonate (Ichthammol) 20% ointment (Fig. S7).

### Imbalance of immune and cytokine inflammation CI patients

To characterize inflammatory responses in CI patients across the dysbiosis clusters, we measured immune cell levels and cytokine responses. First, levels of granulocytes (neutrophils, eosinophils, and basophils) were measured from blood acquired from CI patients (no skin wounding; P2-P5) and healthy controls (P1). There was a statistically significant elevation in neutrophil count for all CI clusters (P2-P5) compared to healthy individuals (P1) (Fig. 5A, left panel). Interestingly, only CI patients from P4 (LI and HI) demonstrated a significant elevation in eosinophil counts (Fig. 5A, center panel), whereas there were no differences in basophils counts among CI patients compared to healthy controls (Fig. 5A, right panel).

**Figure 5.**
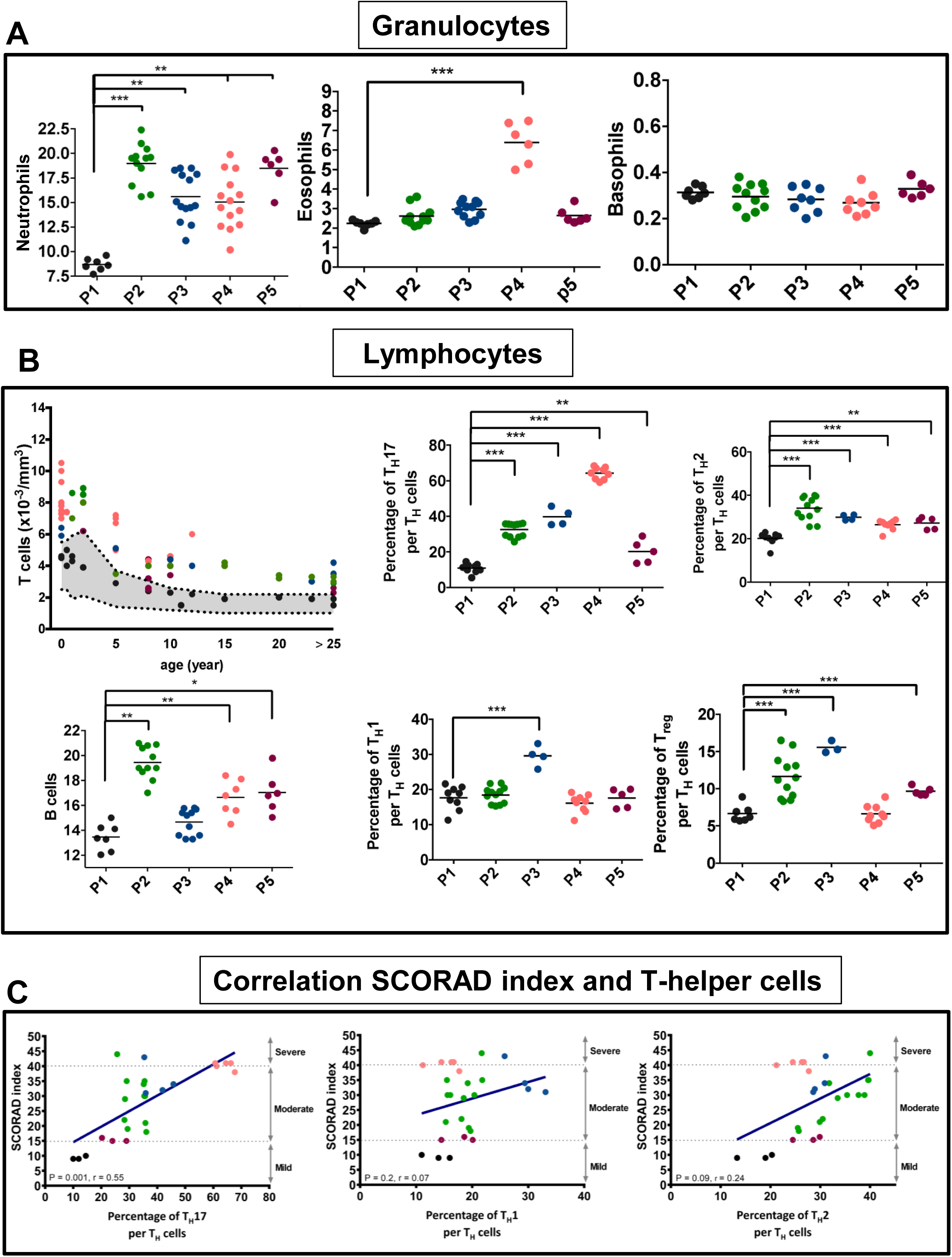
Immunotyping profile of each CI dysbiosis cluster. A. Cell counts of granulocytes including neutrophils (left), eosinophils (center), and basophils (right) from healthy controls (P1) and CI patients (P2-P5) as determined by flow cytometry. Results are expressed in dot plot with each horizontal bar representing the mean of the group with statistical significance as *\*p* ≤ 0.05, *\*\*p* ≤ 0.01, *\*\*\*p* ≤ 0.001, *\*\*\*\*p* ≤ 0.0001. B. Cell counts and relative percentages of lymphocytes including T, B, T_H_17, TH1, TH2, and Treg cells as determined by flow cytometry for healthy controls (P1) and CI patients (P2-P5). Results are expressed in dot plot with each horizontal bar representing the mean of the group with statistical significance as *\*p* ≤ 0.05, *\*\*p* ≤ 0.01, *\*\*\*p* ≤ 0.001, *\*\*\*\*p* ≤ 0.0001. C. Correlation between SCORAD index with percentages of Th17, TH1, and TH2 cells in CI patients. The solid regression line r=0.8195 for Th17 vs. SCORAD; r=0.102 for Th1vs. SCORAD; r=0.423 for Th2 vs. SCORAD. Results are expressed in dot plot.

Similarly, levels of T- and B- cells were quantified from blood from CI and healthy patients (Fig. 5B). Percentage of T_H_2^+^/CD4^+^- and T_H_17^+^/CD4^+^- cells are significantly increased in CI patients (P2, P3, P4, and P5) compared to healthy controls (Fig. 5B, upper right panels). Total T_H_1^+^/ CD4^+^- cells are significantly increased in P3 CI patients, while Treg^+^/CD4^+^ cells are significantly increased in P2, P3, and P5 CI patients compared to healthy controls (P1) (Fig. 5B, bottom right panels). B cells were significantly induced in P2, P4, and P5 CI patients compared to P1 (Fig. 5B, bottom left panel). Importantly, CI patients were clinically subdivided into severe- (score > 40), moderate- (score = 15-40), and mild- (<15) SCORAD index [24], a clinical tool for scoring the severity of atopic dermatitis/eczema; here it is applied to CI. SCORAD index in our CI cohort positively correlated with T_H_17 and T_H_2 (Fig. 5C).

Next, we measured circulating cytokine mRNA levels of T_H_1 (IFN-𝛄, TNF-𝛂), T_H_2 (IL-4, IL-5, IL-13, CCL18), T_H_17 (IL-1𝛃, IL-6, IL-17A, IL-17F, IL-22, CCL20) and Treg (IL-10, TGF-𝛃) cytokines from peripheral-blood leukocytes (Fig. 6). CI patients (P2-P5) were generally found to have higher mRNA levels of T_H_17 cytokines compared with healthy control (P1) (Fig. 6A), except for notable insignificance in IL-22 in P3 and IL-1𝛃, IL-6, IL-17A, IL-17F, CCL20 in P5 compared with healthy control (P1) in two-way ANOVA analysis (Fig. S8A). Similarly, the mRNA level of IL-4, IL-5, IL-13 and CCL18 of T_H_2 was induced in CI patients (P2, P3 and P5) (Fig. 6B), but mRNA levels of IL-13 were not significantly elevated in P5 compared to P1 nor for any of the T_H_2 cytokines in P4 compared to P1 (Fig. S8B). In addition, an increase in the mRNA level of T_H_1 cytokines IFN-𝛄 and TNF-𝛂 was detected in P3 compared to P1 (Fig. 6C), and P3 levels of IFN-𝛄 and TNF-𝛂 were significantly higher than the other dysbiosis clusters (P1, P2, P4, and P5) (Fig. S8C). The mRNA levels of IL-10 and TGF-𝛃 Treg cytokines were increased in P2, P3, and P5 compared to P1, but not P4 (Fig. 6D). These cytokine elevations were significantly highest in P3 when compared with P1, P2, and P5 (Fig. S8D).

**Figure 6.**
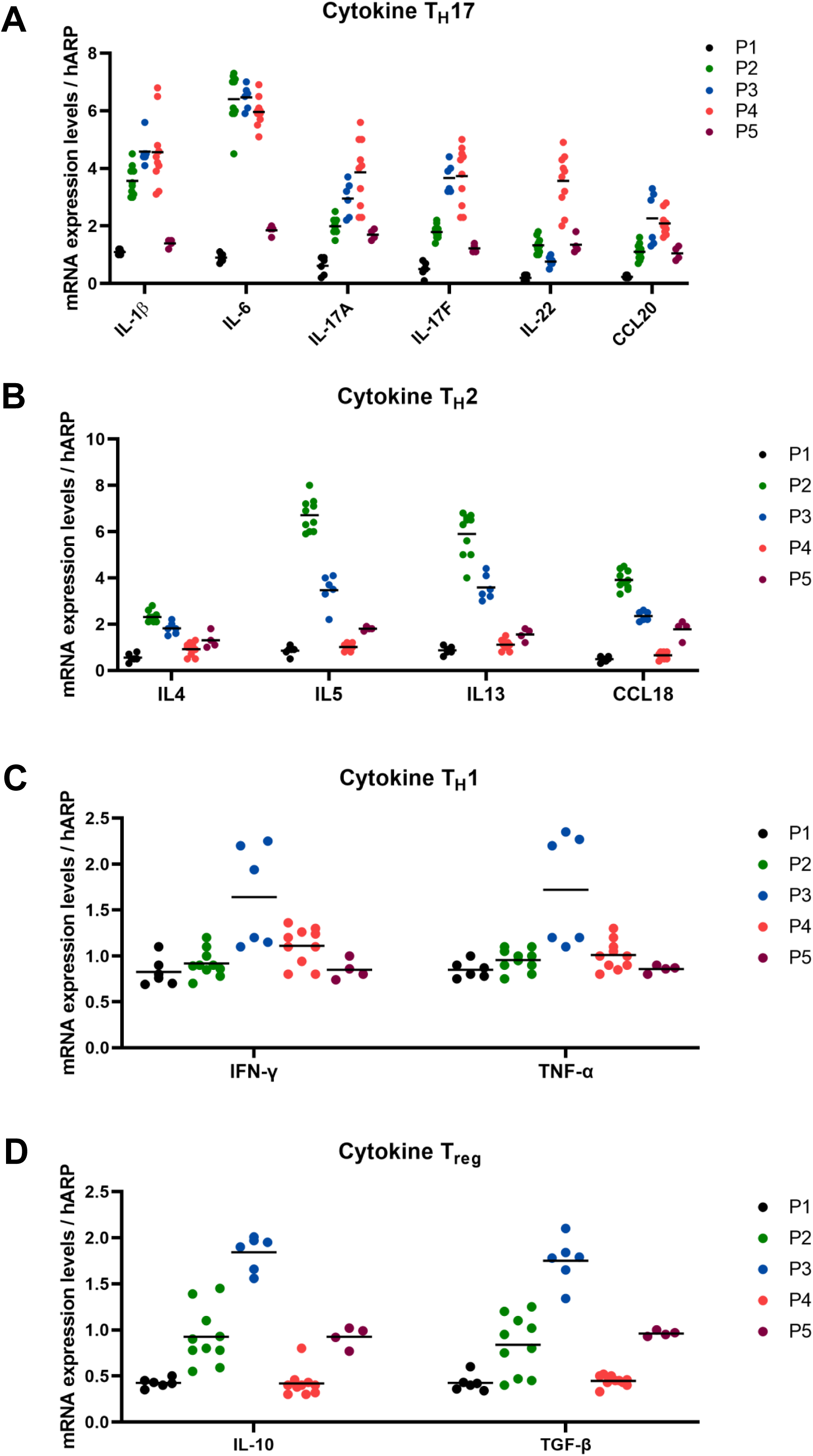
Immunocytokines profiles of CI patients analyzed across the CI dysbiosis clusters. A-D. mRNA expression levels of Th17 (A), Th2 (B), Th1 (C), and Treg (D) cytokines across P1-P5 dysbiosis clusters. Results are expressed in dot plot with each horizontal bar representing the mean of the group.

To further analyze the signaling pathways in CI patients, we evaluated the levels of phosphorylated STAT3 Tyr705 (pY705) as a marker of Janus kinase (JAK) signaling activity in peripheral blood mononuclear cells (PBMCs) (Figs. 7 and S9). Levels of STAT3 pY705 were significantly elevated in CI patients as a group compared to healthy controls (Fig. 7A). Analysis by dysbiosis cluster revealed profound elevations in STAT3 pY705 in P2, P3, and P4 compared with P1, but not in P5 (Fig. 7B).

**Figure 7.**
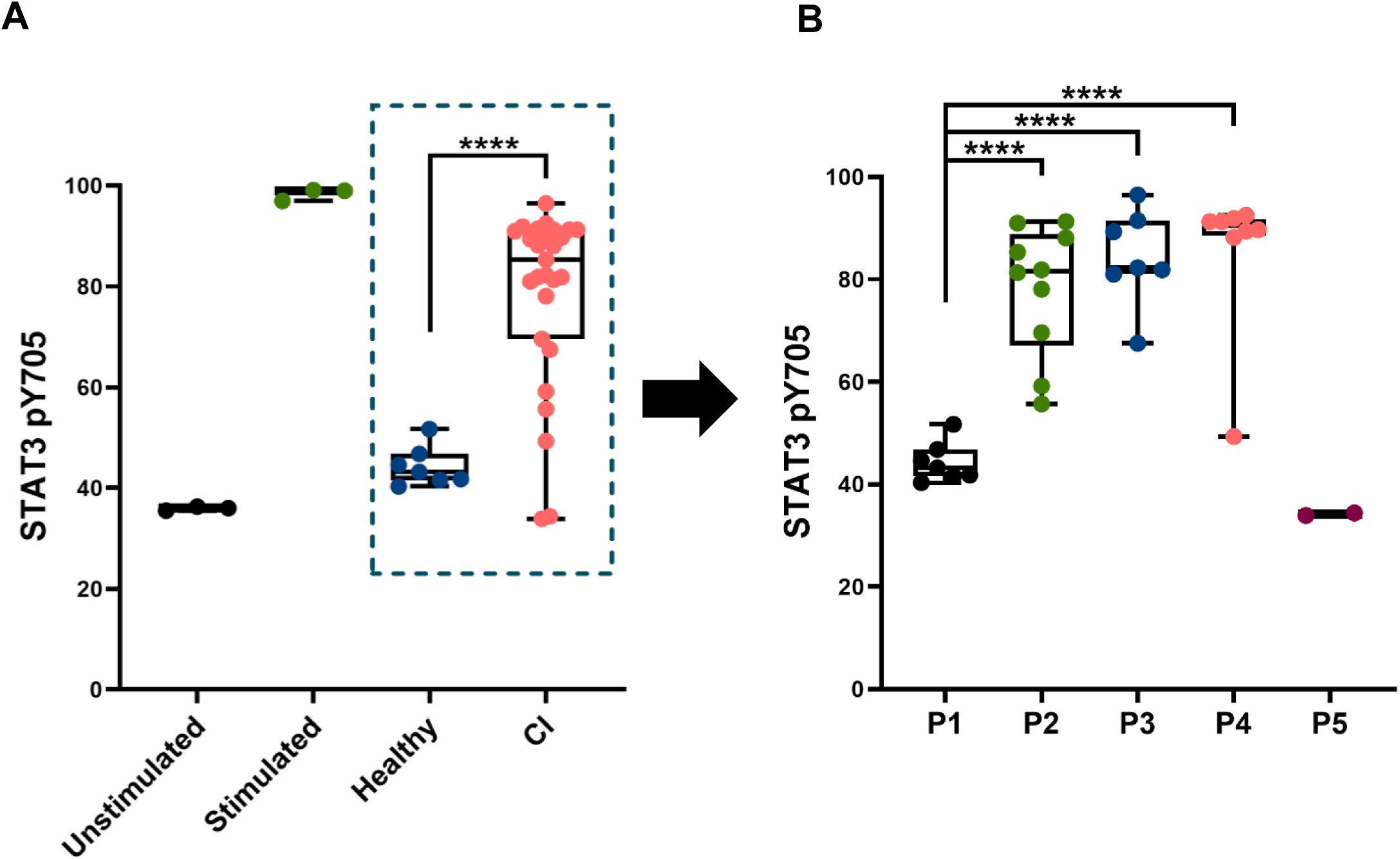
STAT3 pY705 levels as a marker for JAK/STAT signaling in CI patients. (A) Dot plot depicts STAT3 pY705 measured by flow cytometry in subcellular PBMCs. Line bar represents mean. Control samples were unstimulated or stimulated for 15 min with 400 μl Combo (cytokines (100 ng/ml), LPS (10 μg/ml; Sigma-Aldrich) or PMA (100 ng/ml; Sigma-Aldrich)). Healthy and CI patient samples were unstimulated by any substances. Results are plotted in dot plot showing median with interquartile range and statistical significance as *\*p* ≤ 0.05, *\*\*p* ≤ 0.01, *\*\*\*p* ≤ 0.001, *\*\*\*\*p* ≤ 0.0001. (B) Dot plot depicts STAT3 pY705 analyzed across the 4 CI dysbiosis clusters (P2, P3, P4, and P5) compared with healthy controls (P1). Results are plotted in dot plot showing median with interquartile range and statistical significance as *\*p* ≤ 0.05, *\*\*p* ≤ 0.01, *\*\*\*p* ≤ 0.001, *\*\*\*\*p* ≤ 0.0001.

Having established that CI patients generally display elevated neutrophils, Th17 cells, pro-inflammatory cytokines (IL-1𝛃, TNF-𝛂, IL-6, IL-10, and IL-17A/F), and activated JAK signaling/STAT3 p705 compared to healthy controls, we next investigated how the immune response is affected by wounding and wound healing in CI patients. Granulocyte and monocyte populations were analyzed by flow cytometry at baseline (0h), 16h, and 24h after skin wounding using a 5 mm punch biopsy (Fig. S10). Gr1^+^ granulocytes were elevated across P1-P5 at 16h after wounding, but by 24h post-wounding all returned closer to baseline except P2 and P3 (Fig. S10B). This suggests prolonged inflammation in P2 and P3 CI patients post-wounding. Similarly, levels of Ly6^+^ monocytes increased across P1-P5 at 16h post-wounding, and remained more elevated among P2, P3, and P5, and less so P4, compared to P1 at 24h post-wounding (Fig. S10C).

To further probe wound healing response in CI patients, we examined specifically the neutrophil, Th17/Treg, and dopamine-2 receptor (D2R) neural antibody counts in CI patients and healthy controls over 24h, 48h, and 7 days, respectively, post-wounding with a 5 mm punch biopsy (Figs. S10D-F). There was a statistically significant higher proportion of neutrophils in CI patient clusters P2, P3, and P4 versus healthy controls (P1) at 24h post-wounding, but not for CI cluster P5 (Fig. S10D). The Th17/Treg count ratio was statistically elevated for all CI patients (P2-P5) compared to healthy controls at 48h post-wounding (Fig. S10E), indicating skin wounding in CI patients leads to a prolonged Th17 inflammatory response. Lastly, we examined D2R neural antibody as a measure of itch and observed statistically significant elevations in D2R levels before wounding as well as 7d post-wounding across CI patients in clusters P2-P5 (Fig. S10F).

## Discussion

Patients with CI can exhibit a range of symptoms, from mild, acute skin itching to chronic viral and bacterial infections of thick, scaly, fissured skin, with risk of severe or recurrent sepsis, and psychophysiological sequelae [25]. Prior to this study, there was limited data to understand the microbiome and immunological signatures accompanying the various ichthyoses, and even less data across races or ethnicities. We note after preparation of this report a microbiome study on 22 CI patients from 4 types (Congenital ichthyosiform erythroderma (CIE), LI, EI, Netherton’s Syndrome (NS)) was published [26]. Using WES we fully characterized the mutations underlying disease in 36 CI patients belonging to 7 CI types with Southeast Asian ethnicity, identifying 20 novel variants. Even with 36 patients and 7 subtypes, we acknowledge limitations in the overall participant size and CI subtypes in our analysis, primarily due to the rarity of CI genetic disorders. Other limitations were access to care for patients in poor, rural Vietnam regions, and delays in data collection and analysis due to the COVID-19 pandemic. We demonstrated that CI patient microbiota clusters into particular dysbiosis/patient groups according to ratios of pathogenic and non-pathogenic bacteria using 16S rRNA sequencing and PCA. These microbiome signatures directly and differentially impacted wound healing and underlying immune cell populations and cytokine responses in flow cytometry and multiplex ELISA experiments. Our detailed microbiota characterization and immune phenotyping in a clinically and genetically well-defined patient cohort provides key disease insights that may improve CI clinical management.

This work centered around the concept of genotype-structurotype-phenotype correlation [27], which first connects genetic variations (genotype) to altered protein structure and protein malfunction (structurotype), then relates those findings to the patient clinical exam and molecular signatures in the microbiome and immune profile (phenotype). Our microbiome analysis used three different skin sites (dry, moist, and sebaceous), two methods of quantification of microorganisms (cfu and OTU), and evaluated for bacteria, viruses, and fungi. Our study revealed marked differences in the prevalence of Actinobacteria, Firmicutes, Proteobacteria, and Bacteroidetes in the three different sites of skin. Similar to other skin disorders [28–30], our data showed low diversity of all phyla in dry, moist, and sebaceous skin sites of CI patients. Dermatologic conditions often share signatures of microbiome alterations [19]; for instance, *S. epidermidis* and *C. acnes* are less abundant than *S. aureus* within cutaneous inflammation in psoriasis patients [8]. Similarly, *S. aureus* is dominant in patients with ADAM17-deficiency related atopic dermatitis [31], while chronic *C. albicans* is often associated with impaired Th17 related genes [32].

PCA analysis of the microbiome data separated the CI patient cohort into 5 dysbiosis clusters (Fig. 4A, B), including the healthy subjects. Division of the 7 CI subtypes into 4 dysbiosis clusters generated insights into differences in the microbiological and immunological features between these diseases. In unwounded skin, most CI patients had lower abundance of *S. epidermidis*, normally a healthy skin commensal, as well as *C. acnes*. For IV (P2) and EI (P3), these CI types had elevated *S. aureus* and *Malassezia* spp. different from healthy controls and other CI types. This suggests IV and EI patients may benefit from a combination of anti-staphylococcal and anti-fungal therapies. Interestingly, LI and HI CI types clustered together (P4), and demonstrated notable decreases in abundance of *S. aureus*, *S. hominis*, *Malassezia* spp., *B. uniformis* and *B. vulgatus*. *Moraxella* species were elevated in all CI clusters except P5 (ARC, SLS), whereas TTD was the only CI type to demonstrate elevated *B. pseudomallei*.

To better understand how the microbiome of CI patients reacts to skin wounding, we used a 5 mm punch biopsy as a model for skin wounding. While other models or approaches could be used for skin wounding, we found the punch method effective in elucidating differences in the microbiome and immune profile in wounded vs non-wounded CI skin. For example, while *S. aureus* abundance was slightly higher than healthy controls for all CI types in unwounded skin, it was substantially increased by skin wounding. There was a concomitant decrease in *S. epidermidis* in wounded CI skin. Importantly, there was a significant delay in wound healing time for CI clusters P2, P3, and P4, and a smaller delay for P5. We attribute this wound healing delay directly to the microbiome alteration because when given oral β-lactam antibiotics, the wound healing time for all CI clusters normalized closer to the healthy control. This data suggests that oral antibiotics, used with appropriate antibiotic stewardship in mind, may be an effective therapy for healing skin wounds in CI patients. Moreover, this data reinforces that injury or wounding in CI patients is a major risk factor for cutaneous *S. aureus* infection, which may correlate with risk of sepsis and neonatal lethality. Newborn CI patients in particular are at increased risk for health care–associated infections if the barrier of the skin is damaged. Interestingly, we note that the IV patients in our cohort who experienced sepsis (Table S1) had either frameshift or nonsense mutations in the N-terminal half of *FLG* whereas those having mutations more C-terminal did not experience sepsis (Fig. 2). This indicates the type of mutation, its gene location, and its impact on protein production or structure can influence the CI disease course. IV is associated with earlier onset and worse severity of atopic dermatitis [33], which in turn is associated with higher *S. aureus* infections [31, 34].

Immune profiling of the CI cohort with unwounded skin revealed several commonalities: all had elevations of neutrophils, normal basophil levels, and elevations of T_H_17 and T_H_2 T-lymphocytes. Differences, however, emerged among the CI clusters. For example, LI and HI patients (P4) were the only CI cluster with elevations in eosinophils. T_H_17 elevations were not uniform either: P4 had the highest percentage of T_H_17 cells, followed by P3>P2>P5. EI and TTD (P3) patients were the only CI cluster to have elevated percentages of T_H_1 cells, whereas P2, P3, and P5 had elevated T_reg_ cells. At the cytokine level, it follows from the T_H_17 elevations that IL-1𝛃, IL-6, IL-17A and IL-17F were also elevated based on mRNA expression levels, more so for P2, P3 and P4 than P5. For P2 and P3, IL-5 and IL-13 were elevated more than IL-4. Interestingly, upon wounding of the CI skin, there is a notable increase in the ratio of T_H_17 cells: T_reg_ cells for at least 48 hours, indicating a prolonged Th17 inflammatory response. Such potent ongoing inflammatory responses may create a continual inflammatory feedback loop limiting the ability of such patients to effectively heal following wounding (Fig. 8).

**Figure 8.**
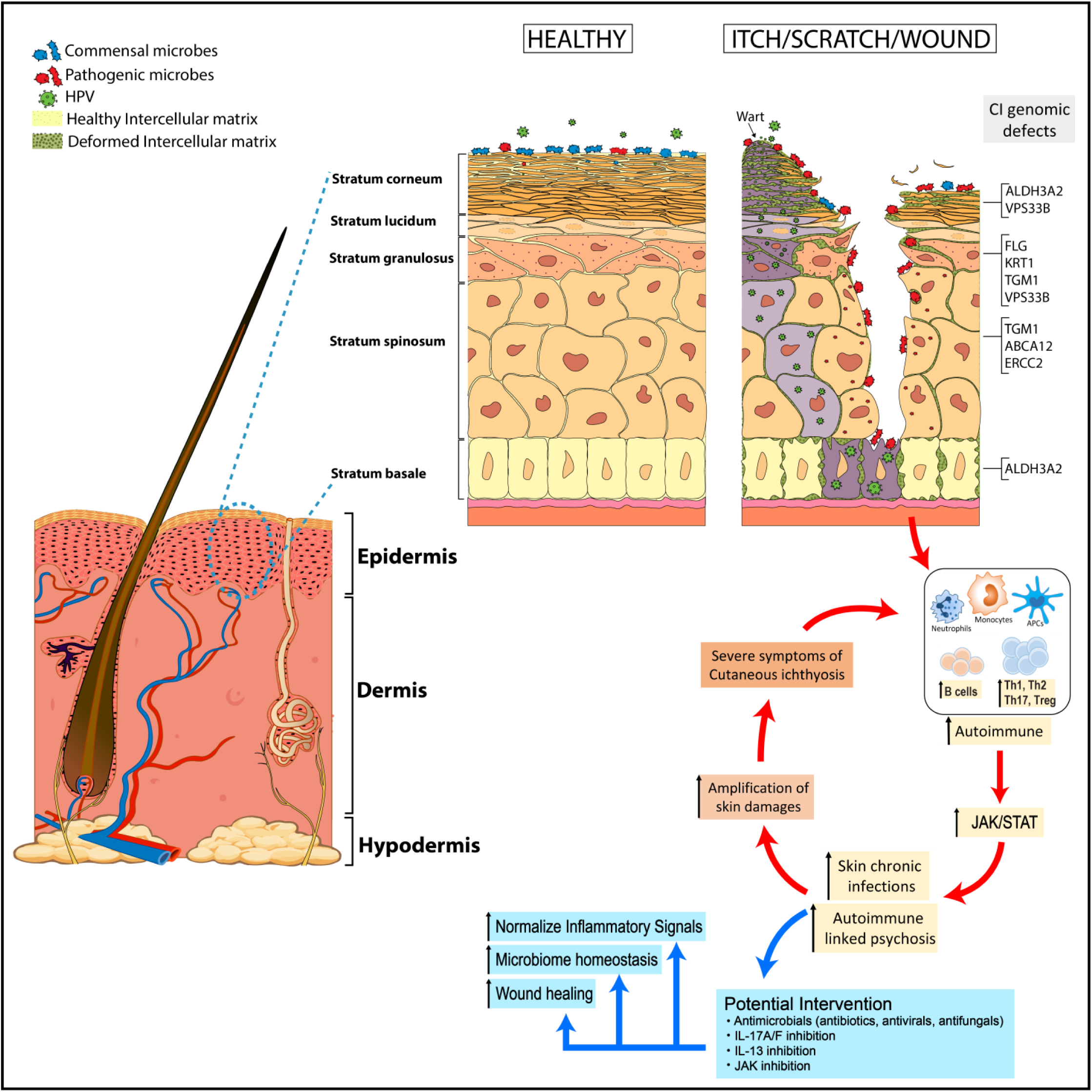
Model of cutaneous ichthyosis related to the barrier change in epidermis layers. The stratum corneum (SC) is the outermost layer of the epidermal barrier. The corneocytes of the SC act as bricks that form a hydrophilic wall that is surrounded by a lipophilic ‘‘mortar’’ made up of lipid lamellae, which fill the extracellular space. Genomics defects underlying CI diseases alter the epidermal barrier at various levels (SC, stratum spinosum (SS), and stratum basale (SB)), enabling pathogenic microorganisms to proliferate, invade, and cause microbial dysbiosis in the skin. In CI patients, as shown in this study, microbial dysbiosis and/or skin wounding amplifies Th17-driven inflammation in the skin, which can be associated with elevated JAK/STAT signaling.

Our data corroborates the heightened Th17 inflammatory response seen previously in CI patients [19–21], which has stimulated investigations of the IL-17A inhibitor secukinumab as therapy for CI. While there are sporadic case reports of secukinumab improving a patient with autosomal recessive congenital ichthyosis harboring an *ABCA12* variant [35], as well as patients with Netherton’s syndrome [36, 37], a double-blind randomized controlled trial evaluating secukinumab in 4 types of CI patients compared to placebo failed to reduce disease severity or reduce Th17-related biomarkers [38]. Consistent with one study from 2019 [20], our data shows all CI dysbiosis clusters with elevated IL-17A and IL-17F. These two cytokines have synergistic/cooperative effects on one another [39], and clinical trial data for bimekizumab, a biologic with dual IL-17A and IL-17F inhibition, has achieved high rates of Psoriasis Area and Severity Index (PASI)-100 clearance [40]. Thus, it is plausible that treatment of CI patients with dual IL-17A/F inhibition will prove more effective than IL-17A inhibition alone.

The recent emergence of JAK inhibitors in dermatology, particularly those Food and Drug Administration-approved for atopic dermatitis (ruxolitinib, upadacitinib, abrocitinib), makes our finding of elevated JAK-STAT signaling in CI patient clusters P2, P3, and P4 intriguing. This suggests that JAK inhibitors, or even TYK2 inhibitors, might be effective in alleviating skin inflammation and barrier defects in many CI patient types. In fact, elevated JAK-STAT signaling activating the nitric oxide synthase (NOS2) pathway in HI has been documented and alleviated using tofacitinib in a 3D model of HI [41]. Moreover, itch was observed in 97.2% of our CI cohort, and JAK inhibitors to date have proved successful in reducing itch [42]. Another common driver of itch, IL-13 [43], was elevated for IV, EI, and TTD patients (P2 and P3), suggesting that the atopic dermatitis biologic tralokinumab, an IL-13 inhibitor, might prove useful in reducing itch and inflammation in some CI patients [44], or alternatively, the IL-4/IL-13 inhibitor dupilumab. Ultimately, the dysbiosis clustering described here suggests that optimized treatment regimens for CI types will have to account for their heterogeneity from a genetic, microbiome, and immunological viewpoint.

## Materials and Methods

### Sample collection for germline sequencing and microbiome

CI patients were recruited to participate in a study approved by the Institutional Review Board of UMP Ho Chi Minh City from December 2013 based on the Newton Researcher Link British Council project in compliance with all ethical regulations. From December 2016 we continued to recruit samples from the Nafosted project, supervised by Dr Nga Nguyen at Dalat University. Written informed consent was obtained from all adult patients and the parents/guardians of all participating children (under 16-year-old). Subjects provided detailed medical history and underwent a physical examination at Children’s Hospital 2, Hung Vuong Maternity Hospital, UMP Medical Centre 3 and City Children’s Hospital, Vietnam. Follow-up schedule was separated in two periods (from 9/2013 to 9/2016, for DNA exome sequencing, and from 9/2018 to 12/2019, for wound healing test).

All samples were separated into 3 blood tubes (BD Biosciences); one EDTA vacutainer for germline DNA isolation, one CPT heparinized vacutainer for immune cell isolation, and one silicon-coated serum tube for cytokine analysis. All sterile Dacron swabs (Fisher Scientific, Sweden) from dry, moist, sebaceous, and lesioned skin were collected and DNA immediately extracted using the MO-BIO PowerSoil DNA isolation kit (MO-BIO Laboratories), otherwise, raw specimens were stored in freeze (−80°C) until isolation of gDNA. Negative control swabs were also collected for each patient sample. Swabs were processed and cultured at the hospital department of microbiology following standard protocol to confirm the common skin bacteria as well as methicillin-resistant *S. aureus* infection processed [45]. All extracted DNA was quantified by NanoDrop and passed quality control testing prior to both WES or 16S-based metagenomic sequencing.

### Clinical whole exome sequencing

Clinical WES was described previously [46]. In brief, genomic DNA was extracted from peripheral blood using QIAamp DNA Blood Mini Kit (#51104, QIAGEN). DNA was subsequently examined for quality using DropSense96, Qubit 2.0, and TapeStation. The ideal range for these parameters were: chemical contaminants A260/A230: 1.50 – 2.50; protein contaminants A260/A280: 1.60 - 2.20; concentration > 1 ng/µL; DNA smear (50% > 1000 bp). Whole exome library preparation and sequencing was performed by Macrogen (South Korea), using Agilent SureSelect Human All Exon V5 (Agilent Technologies) on a NovaSeq 6000 Sequencing System (Illumina). Using paired FASTQ reads, FASTQC was used to obtain diagnostics such as Phred-score distribution along the reads, GC content distribution, read-length distribution, and sequence duplication level. After that, Trimmomatic was used for removing contaminated sequencing adapters; removing leading and trailing low quality or N bases below quality 3; scanning the read with a 4-base wide sliding window, cutting when the average quality per base drops below 15; and dropping reads below a length of 36. Pre-processed read pairs were then mapped to hg19 reference genomes (from UCSC) by BWA-mem. Additional processing included: MarkDuplicates by PICARD, BaseQualityScoreRecalibration by GATK. VCF files were generated with GATK HaplotypeCaller; filtrated by GATK VariantFiltration, SNP (QD < 2.0, FS > 60.0, MQ < 40.0, MQRankSum < −12.5, ReadPosRankSum < −8.0) and INDEL (QD < 2.0, FS > 200.0, ReadPosRankSum < −20.0), respectively. Finally, ANNOVAR was used to intersect variant annotations from UCSC RefSeq, dbSNP 150, gnomAD, ESP6500, ExAC, 1000G, dbNSFPv3.5. The variant determination and classification are based on VarSome’s bioinformatics [47] that match the guideline of ACMG 2015 and all variants confirmed by Sanger sequencing.

### Microbiological investigations

DNA was isolated from frozen archived swab specimens with automatic platform Cobas4600 (Roche); then were loaded to array with AccuFill system. Specific strain was confirmed by blood cultures, bacterial cultures, and by multiplex PCR from TaqMan™ Comprehensive Microbiota Control (Thermo Fisher Scientific, Korea), and Antibiotic resistance genes (included sulfonamide resistance genes (sul1 and sul2), trimethoprim resistance genes (dfrA1, dfrA5), beta-lactam resistance genes (ampC, blaTEM, blaSHV and blaPSE-1) and tetracycline resistance genes (tet(B), and tet(M)) by TaqMan Assays (Thermo Fisher Scientific, Korea). Broad targeted genotyping Takara PCR-Realtime detection assay were customed by *Cutibacterium acnes*; *Lawsonella clevelandensis*; *Bacteroides uniformis*; *Bacteroides vulgatus*; *Staphylococcus capitis*; *Staphylococcus aureus*; *Staphylococcus hominis*; *Staphycococcus epidermidis*; *Morazella osloensis*; *Burkholderia pseudomallei*; *Malassezia* spp. Probes (SmartChip® Probe qPCR Master Mix) and TB Green® Advantage® qPCR Premix (Takara, Japan). Probe and primer sets were listed in Table S4.

### Immunological investigations

One mL of whole blood was drawn into a TruCulture^®^ (Myriad RBM) tube containing TruCulture® media. Samples were incubated upright in a dry heat block at 37°C for 30 h. Supernatant and cells were isolated and frozen for cytokines. T, B, subsets were counted by 6-color TBNK BD TruCount (BD Biosciences) from patients and three or four age-matched healthy controls. mRNA cytokine levels were measured in cell-culture supernatants 2 hours after *in vitro* stimulations by qPCR (Thermo Fisher).

### 16S rRNA sequencing and analysis

DNA isolation and library preparation to generate shot-gun metagenomic sequence data from microbiome followed a Macrogen protocol. The V1-V3 region of the 16S rRNA gene was amplified with previously described primer sets [2, 48]. ITS1 amplicons used are as previously reported [49]. The amplicon sequence data produced 15 million to 50 million 2 x 125-bp reads on an Illumina HiSeq platform (Macrogen). Raw sequence data were proceeded by Mothur pipeline to remove primers and barcodes. Chimeric reads from PCR artifacts were identified and trimmed by VSEARCH in Mothur. Secondary clusters were recruited into primary clusters. Noise sequences in clusters of size “a” or below were removed. The reads were clustered using a greedy algorithm into OTU clusters (including OTUs, Chao1, Shannon and Simpson index) at a user-specified OTU cutoff as previously described ([49]. Taxonomic assignment and diversity statistics were analyzed and visualized by Quantitative Insights into Microbial Ecology (QIIME). The qualified data then were classified to genus level using the k-nearest neighbor classifier against an updated custom ITS1 database. Phylotype for both 16S and ITS1 amplicon was analyzed to Mothur SOP. Quality processed reads not matching hg19 human reference were mapped against a database of 2,349 bacterial, 389 fungal, 4,695 viral, and 67 archaeal reference genomes using Bowtie 2 (version 2.3.2)–very-sensitive parameter [50]. Read hit counts were normalized by genome size. To reduce the effect of low abundance misclassifications, we used a genome coverage cutoff of ≥1 for relative abundance and diversity calculations. Metagenomic reads not mapping to bacterial, fungal, or archaeal reference genomes were exported using the parameter of Bowtie 2 for de *novo* assembly.

Taxonomic classification of papillomaviruses is based on nucleotide similarity of the L1 gene51. The family *Papillomaviridae* contains 49 genera (with 5 genera representing human papillomaviruses), each of which is further divided into several species. To be designated as a new type, a single papillomavirus type cannot share >90% similarity to any other known papillomavirus type in the L1 sequence. Papillomavirus types within a species share 71–89% nucleotide identity within the L1 gene and members of the same genus share >60% L1 sequence identity.

### STAT3 signaling analysis

We collect PMDC cells and extracted protein lysate for anti-STAT3 (Cell Signaling Technology, Cat#30835), anti-pSTAT3 (pY705, Cell Signaling Technology, Cat#9145); anti-FAK1. We used the inhibitors of FAK inhibitor 14 (Sigma-Aldrich) and Stattic (Sigma-Aldrich). The drugs Stattic were dissolved in dimethylsulfoxide (DMSO), while FAK inhibitor 14 was dissolved in water at the desired concentrations and stored at −20 °C. The high throughput subtypes of cells via flow cytometry towards detecting the STAT3 and STAT3 pY705 were followed the previous procedure [51, 52].

### R analysis

All statistical analyses were performed using R software. Data are represented as mean ± S.E.M. unless otherwise indicated. Spearman correlations of non-zero values were used for all correlation coefficients. Differences in OTU abundance between two groups were analyzed using White’s non-parametric *t*-test. For all boxplots, center lines represent the median while lower and upper box limits represent the first and third quartiles, respectively (interquartile range), whiskers represent the maximal values up to 1.5 times interquartile range, and all values beyond this range are defined as outliers. The non-parametric Wilcoxon rank-sum test was used to determine statistically significant differences between microbial populations. Unless otherwise indicated, P values were adjusted for multiple comparisons using the Bonferroni or FDR correction (by applying the adjust function in R using method = ‘bonferroni’ or ‘fdr’). Statistical significance was ascribed to an alpha level of the adjusted P ≤ 0.05. Similarity between samples was assessed using the Yue–Clayton theta similarity index with relative abundances of HPV species. The theta coefficient assesses the similarity between two samples based on (i) number of features in common between two samples, and (ii) their relative abundances, with θ = 0 indicating totally dissimilar communities and θ = 1 identical communities.

### Data availability statement

The authors declare that all other data supporting the findings of this study are available within the paper and its supplementary information files.

## Acknowledgements and funding

We thank Dr. Mahmoud Ghannoum for critical review of this manuscript. This study was primarily supported by a Vietnamese Nafosted grant (#106-YS.01-2016.39) (to **H-N.N**), Newton Researcher links (#Newton.01.9.2014-2016) (**C-B.B**), City’s Children Hospital (#CCH-Microbiome2017) (to C-B.B), and the Kenichi Arai research fellowship (#KIA092018) (to **C-B.B**). This work was also supported by grants from the National Institute of Arthritis and Musculoskeletal and Skin Diseases of the National Institutes of Health under Award Numbers K08AR070290, R03AR076484, and R01AR079428 (to **C.G.B**). The content is solely the responsibility of the authors and does not necessarily represent the official views of the National Institutes of Health. We would like to thank all Newton Researcher Links from ImmunoTechniques, Cho Ray’s Hospital, Children Hospital 1, Children Hospital 2, and University Medical Centre. List of doctors and researchers: Than Duc Dung, Quynh Anh Phan, Thong Van Nguyen, Lien Anh Nguyen Phan, Hiep Minh Nguyen, Nguyen Thi Thanh Ha, Vinh Nguyen, Edel O’Toole, David Kelsell, Dieu-Thuong Thi Trinh, Nam Tran Nguyen and Minh Tien Nguyen and Dr. Leigh Ann Jones.

## Author contributions

**M.H**., **H-N.N**.: Conceptualization, Investigation, Writing-Review & Editing, Formal Analysis, Visualization; **M.V.H**., **T.T.T.B**.: Clinical Diagnosis, Investigation & Management; **T.H.T.D**. and **V.H.T.M**.: Immune Investigation and Analysis; **B-Q.V**., **D.B**.: Whole Exome Sequencing Analysis. **M.H**. and **B-Q.V**.: Microbiome Analysis. **S.A.E.**: Visualization; **Newton Researcher Links**: Investigation, Resources; **C.G.B.**: Conceptualization, Validation, Visualization, Writing-Review & Editing, Project Administration, Funding acquisition; **C-B. B.**: Conceptualization, Validation, Formal analysis, Investigation, Resources, Writing-Original Draft, Writing-Review & Editing, Visualization, Supervision, Project administration, Funding acquisition.

## Supplemental Figure Legends

**Figure S1.**
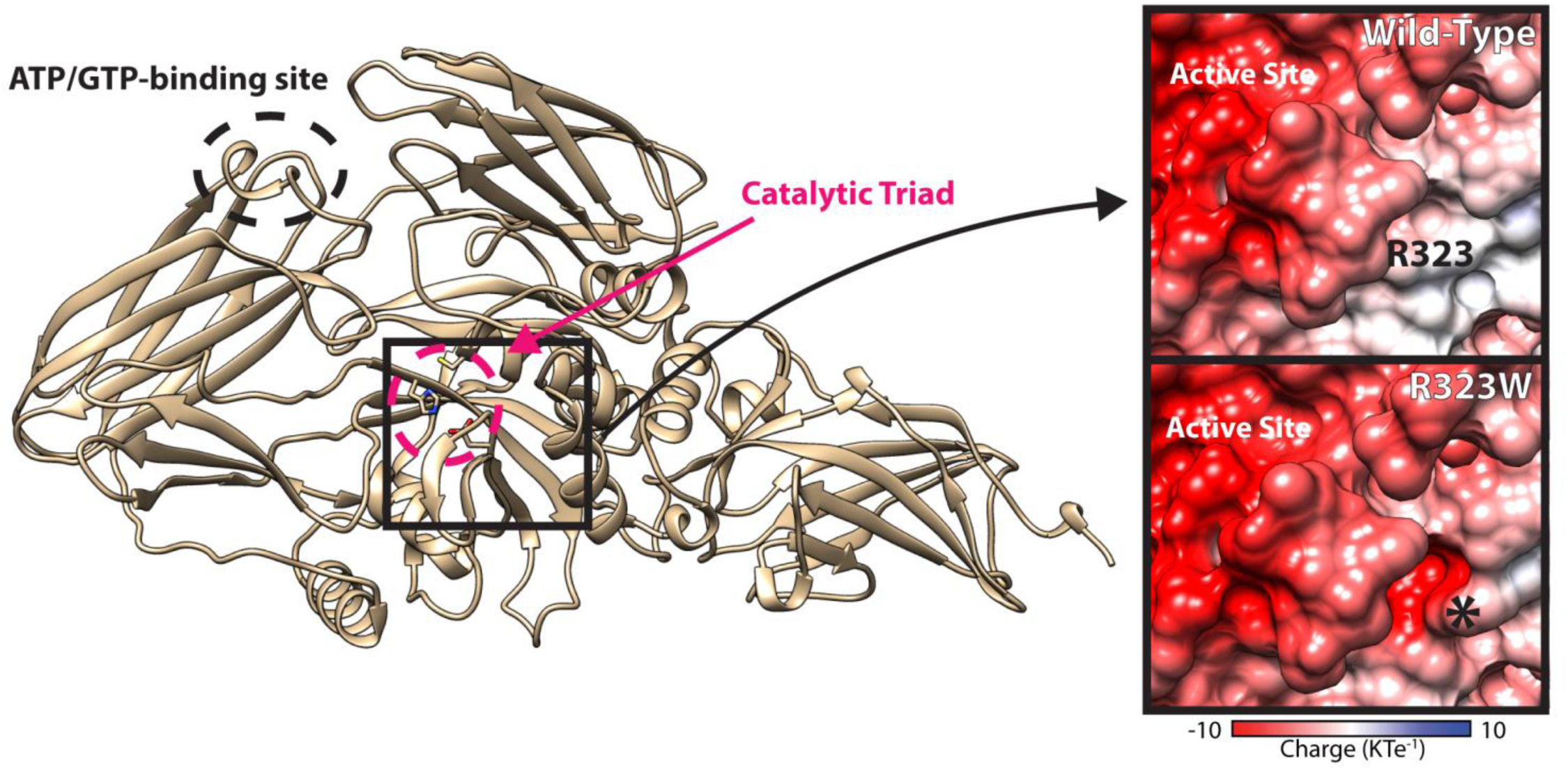
Structural basis of TGM1 mutations in lamellar ichthyosis. A structural model of TGM1 was produced based on coagulation factor XIII (Protein Data Bank (PDB) Accession Code 1F13, which had a higher sequence identity and better GMQE score than the transglutaminase 2 structure). The TGM1 model overlays well with the TGM2 structure (PDB 3LY6) with the catalytic triad (C377, H436, D459, within red circle) well-conserved. R323 is highly conserved in the transglutaminase family and the R323W mutation increases the acidity of the protein surface in a pocket very close to the active site, which is itself very acidic (Herman et al., 2009). TGM1 is cleaved from a low activity to high activity form between S92 and R93, thus the R93Q mutation disrupts this process (Kim et al., 1995).

**Figure S2.**
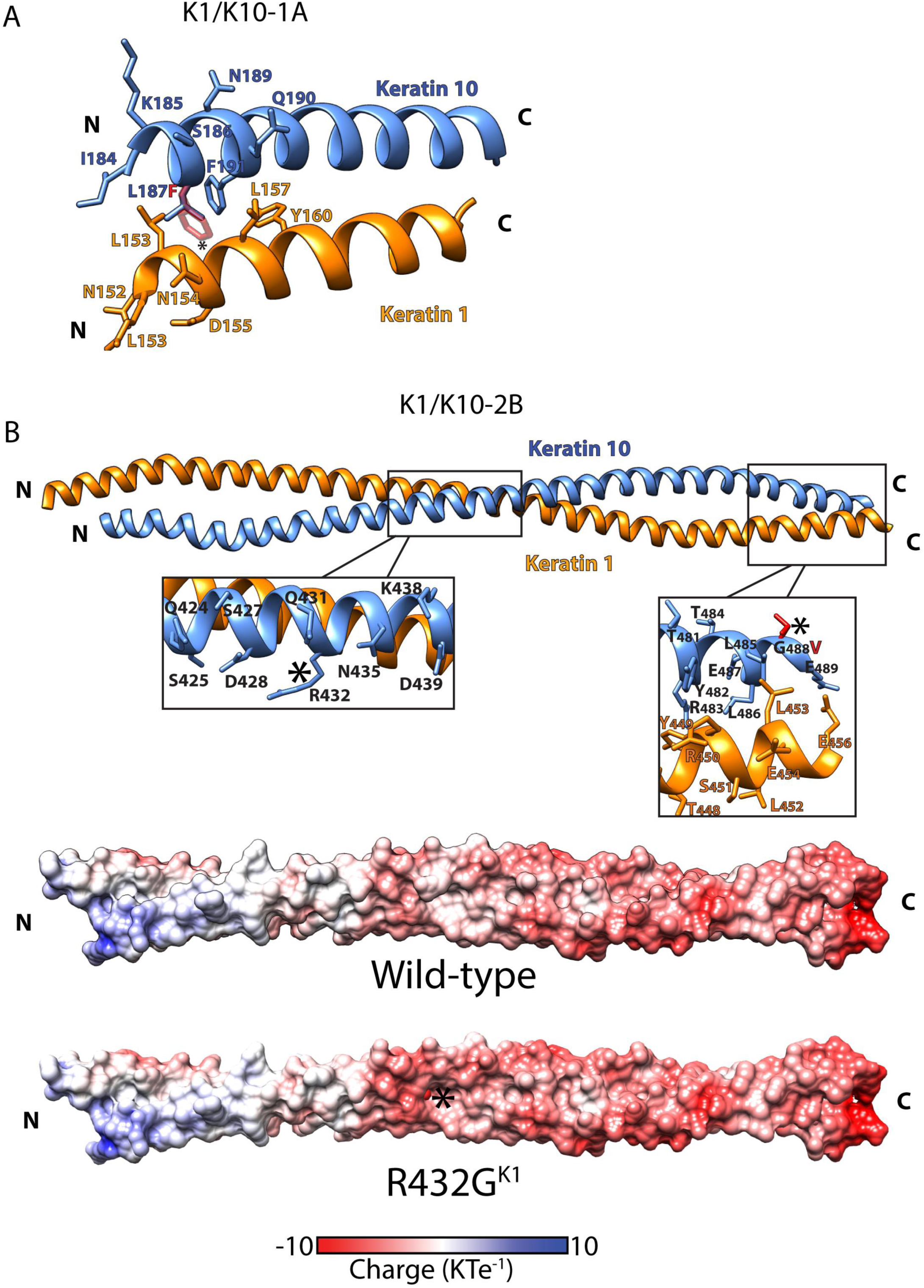
Structural basis for keratin 1/keratin 10 mutations in epidermolytic ichthyosis. (A) A structural model of the K1/K10 helix 1A heterocomplex was produced using the K5/K14 helix 2B crystal structure (PDB Code 3TNU) as a template based on GMQE score. L187F mutation is at the N-terminus of the 1A domain, specifically located at the heterodimer interface. The mutation likely destabilizes the heterodimer due to increased, bulky size of the amino acid at the mutated position causing steric clashes with the K1 backbone as well as the side chains of L153^K1^, L157^K1^, and F191^K10^. (B) The published crystal structure of the K1/K10 helix 2B heterocomplex (PDB Code 4ZRY) was used to investigate the effects of missense mutations in this keratin region. The R432G mutation eliminates a surface-exposed basic residue, thereby reducing positive surface charge and increasing the acidity of the region and likely disrupting an interaction with another keratin molecule required for mature filament assembly. The G488V mutation occurs at the C-terminus in the highly conserved TYR*LLEGE motif known to be critical for intermolecular interactions (Wilson et al., 1992; Lomakin et al., 2020). The G488V mutation is thus likely to disrupt higher order filament assembly.

**Figure S3.**
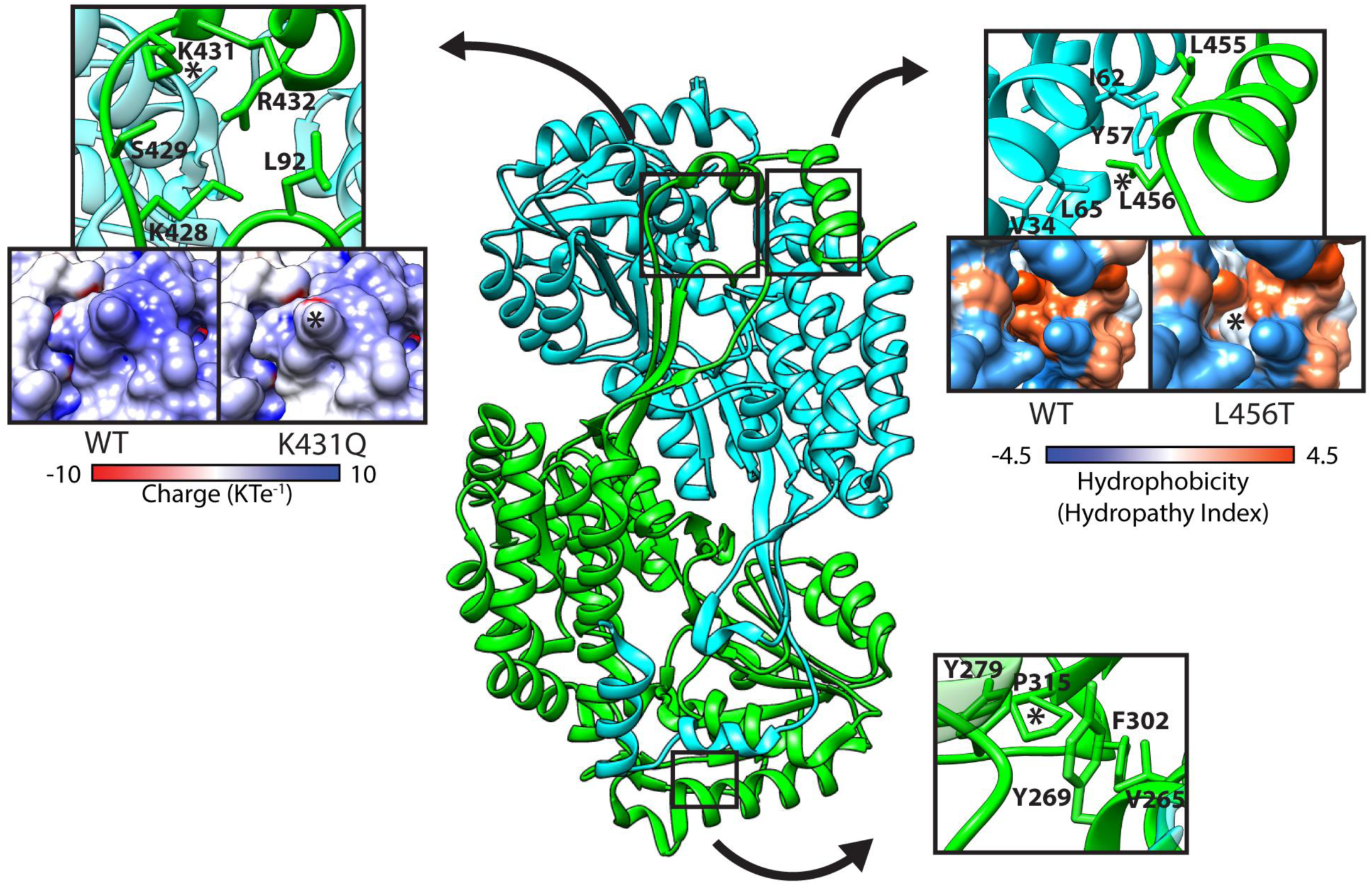
Structural basis for fatty aldehyde dehydrogenase mutations in Sjögren-Larsson syndrome. *ALDH3A2* encodes the membrane-bound fatty aldehyde dehydrogenase (FALDH), which converts long-chain fatty aldehydes to fatty acids. The published x-ray crystal structure of FALDH (PDB 4QGK) was used to investigate missense mutations in *ALDH3A2*. The P315S mutation is predicted to destabilize the folding of the protein as it removes a proline in a loop, changing the orientation of the loop. Furthermore, this mutation introduces a polar serine into an otherwise hydrophobic region buried in the protein, resulting in entropic destabilization of this region. The L456T mutation, which was observed in the same patient with the P315S mutation, also causes disruption of hydrophobic interactions. These two mutations occurring concomitantly destabilize the protein structure to reduce its catalytic activity. The L456T mutation is also significant because it occurs in a helix thought to act as a gatekeeper for substrates. Another individual disease-causing mutation, K431Q, reduces the positive charge at the surface of the enzyme. The charged residues in this region are conserved in the FALDH protein family and have the function of binding the negatively charged head groups of the lipid bilayer. Consequently, the charge reduction caused by the K431Q mutation may reduce substrate affinity and thus impede enzyme activity (Keller et al., 2014).

**Figure S4.**
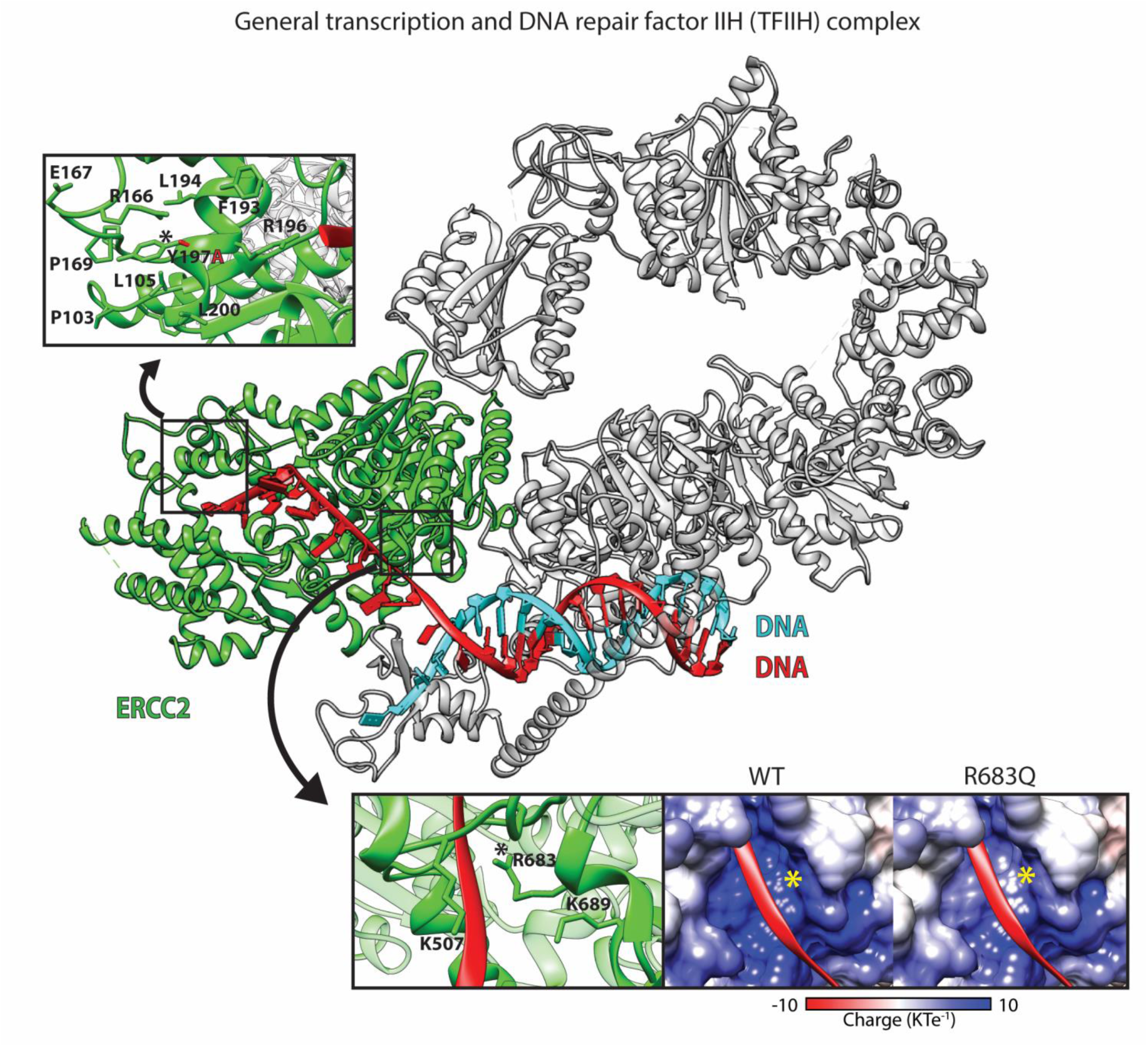
Structural basis for TTD mutations in trichothiodystrophy. *ERCC2* encodes the excision repair protein TTD, which makes up one subunit in the general transcription and DNA repair factor IIH (TFIIH) complex. TTD has helicase and ATP hydrolysis activity, making it an important part of the TFIIH complex (Yan et al). The structure of DNA-bound TFIIH, including TTD, has been determined by cryo-EM (PDB 6RO4) and was used to model the disease-causing mutations Y197A and R683Q. The Y197A mutation may alter the folding of the protein as it significantly decreases residue size and eliminates some hydrophobic interactions (with P169, L105). This may have a noticeable impact on protein function because it occurs one residue away from R196, which directly interacts with the DNA substrate. Similarly, R683 interacts with the DNA backbone. Elimination of the positive arginine charge is expected to reduce the enzyme interaction with the negatively-charged DNA molecule.

**Figure S5.**
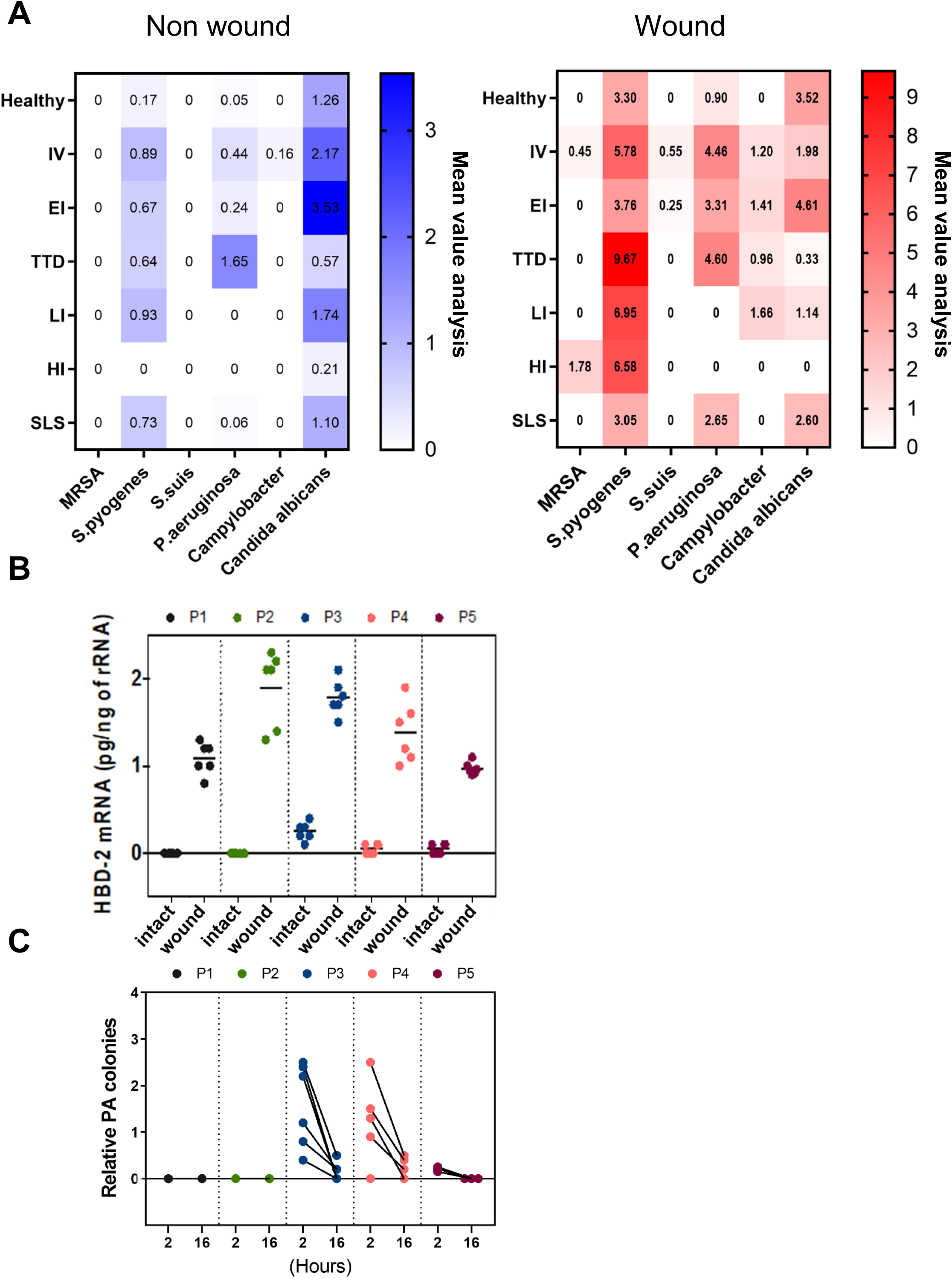
Microbiota and antimicrobial peptide changes in wounded CI skin compared to non-wounded skin. A. Dynamic changes in abundance of select microbial community species in non-wounded (left) and wounded (right) skin of representative CI patients (IV, EI, TD, LI, HI, SLS) compared with healthy controls. B. Level of innate antibacterial β-defensin 2 (mRNA HBD-2) in wounded CI skin (P2, P3, P4, and P5) compared to wounded skin from healthy controls (P1). Results are expressed in dot plot with each horizontal bar representing the mean of the group. C. Measurement of relative colonies of *Pseudomonas* aeruginosa across the 5 CI dysbiosis clusters (P1-P5) for 16 hours after skin wounding.

**Figure S6.**
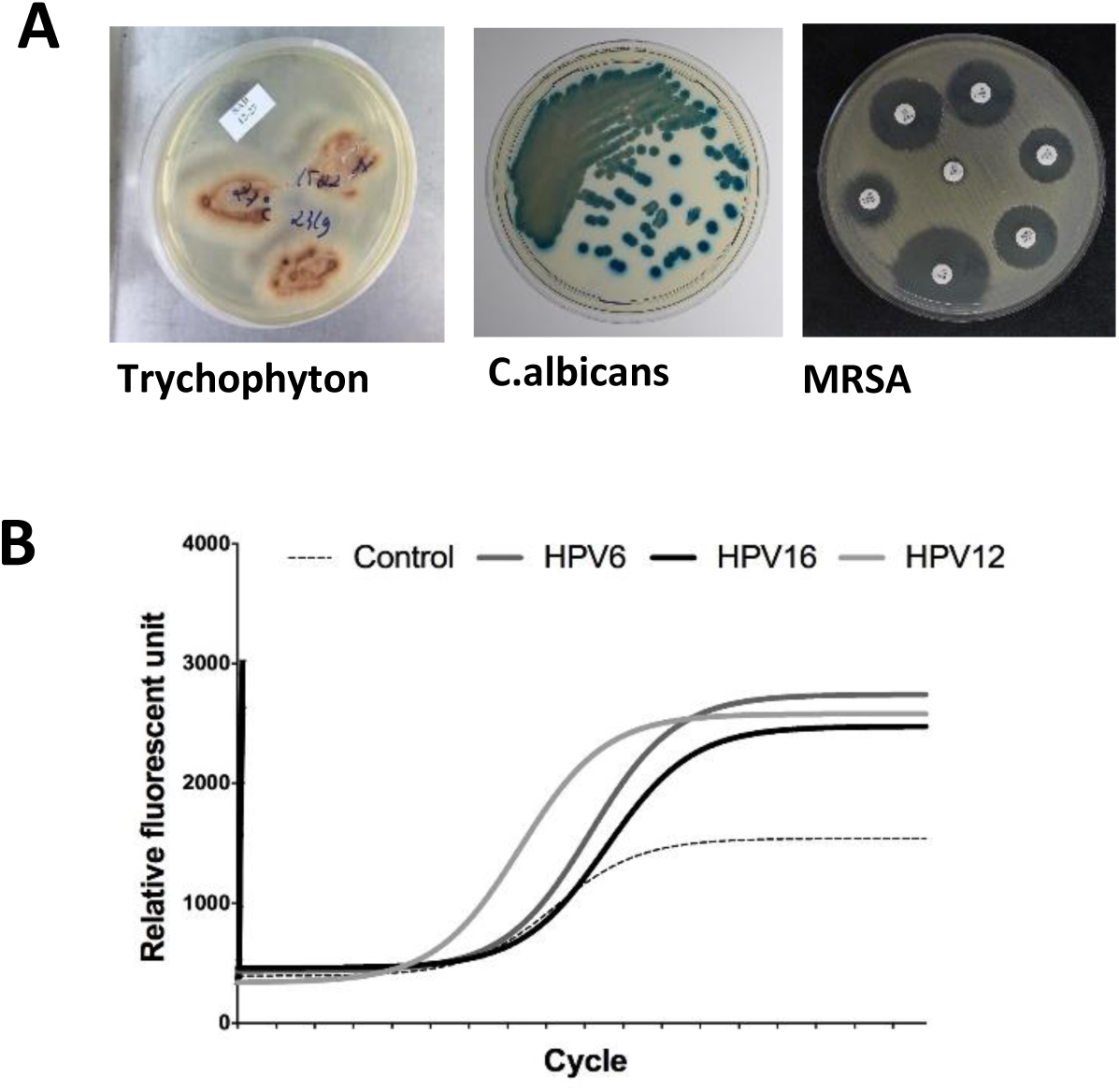
Presence of microbial and viral species in CI patients. A. Bacterial culture of Trychophyton, *C. albicans* and MRSA of CI patients. Samples collected from IV2 infected by Trychophyton and *C. albicans* and from HI6 infected with MRSA, which caused septic shock. B. Detection of human papillomavirus (HPV) subtypes 6, 16, and 12 for TTD2 patient.

**Figure S7.**
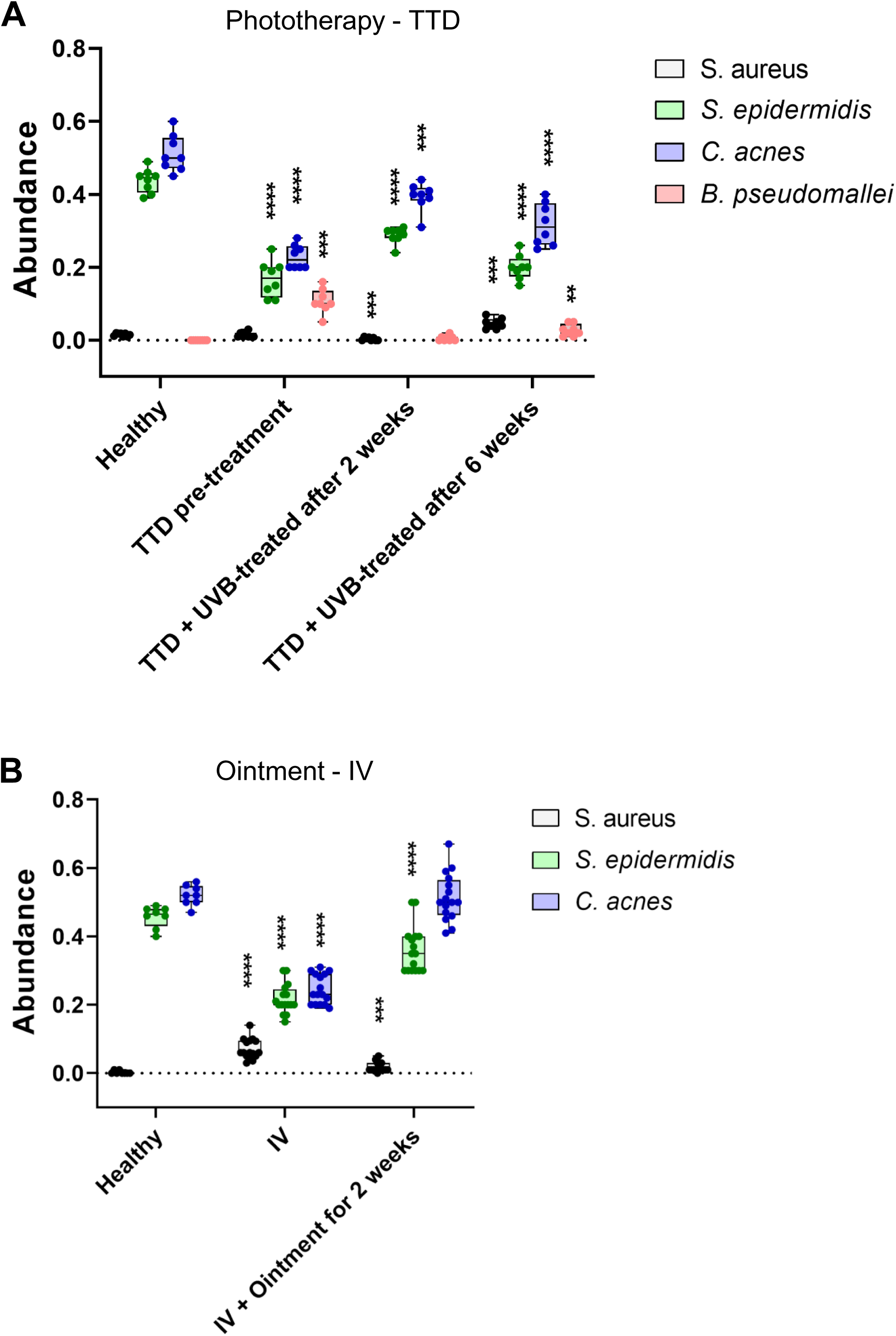
Restoration of CI patient microbiome homeostasis with treatments of TTD and IV. Microbiome changes during therapeutic treatment for TTD and IV subtypes of cutaneous ichthyosis were examined. Participants who received antibiotics or experienced side effects after treatment were excluded. Eight healthy control samples were collected from intact skin on the face, arm, and leg. Relative abundance of *S. aureus*, *S. epidermis*, *C. acnes*, and *B. pseudomallei* colonization was examined with microbiological real-time PCR of the 16S-rRNA test. A. Three TTD patients received narrow-band UVB phototherapy treatment in a mean dose of 25.5 J/cm2 twice weekly for 6 weeks (8 focal lesions on face-nose, forehead, and cheek were collected). Specimens were analyzed from TTD patients prior to treatment, after 2 weeks phototherapy, and after 6 weeks phototherapy to evaluate the targeted species abundance. Phototherapy increased commensal organisms *S. epidermidis* and *C. acnes* closer to healthy levels, while decreasing *B. pseudomallei*. Relapse of skin lesions on TTD patients occurred 6 months after completing phototherapy. Results are expressed in dot plot with median and interquartile range with statistical significance as *\*p* ≤ 0.05, *\*\*p* ≤ 0.01, *\*\*\*p* ≤ 0.001, *\*\*\*\*p* ≤ 0.0001. B. Twelve IV patients (16 focal lesions on leg/arm and on face were collected) applied once daily ointment treatment (ammonium bituminosulfonate (Ichthammol) 20% with zinc oxide (Ichthopaste® bandage)) for 2 weeks along with oral supplemental 1000mcg Vitamin B12. Specimens were taken from IV patients prior to treatment and 2 weeks after treatment to evaluate the targeted species abundance. Topical ointment therapy after 2 weeks reduced *S. aureus* in IV patients and restored *S. epidermidis* and *C. acnes* abundance to levels close to healthy controls. Results are expressed in dot plot with median and interquartile range with statistical significance as *\*p* ≤ 0.05, *\*\*p* ≤ 0.01, *\*\*\*p* ≤ 0.001, *\*\*\*\*p* ≤ 0.0001.

**Figure S8.**
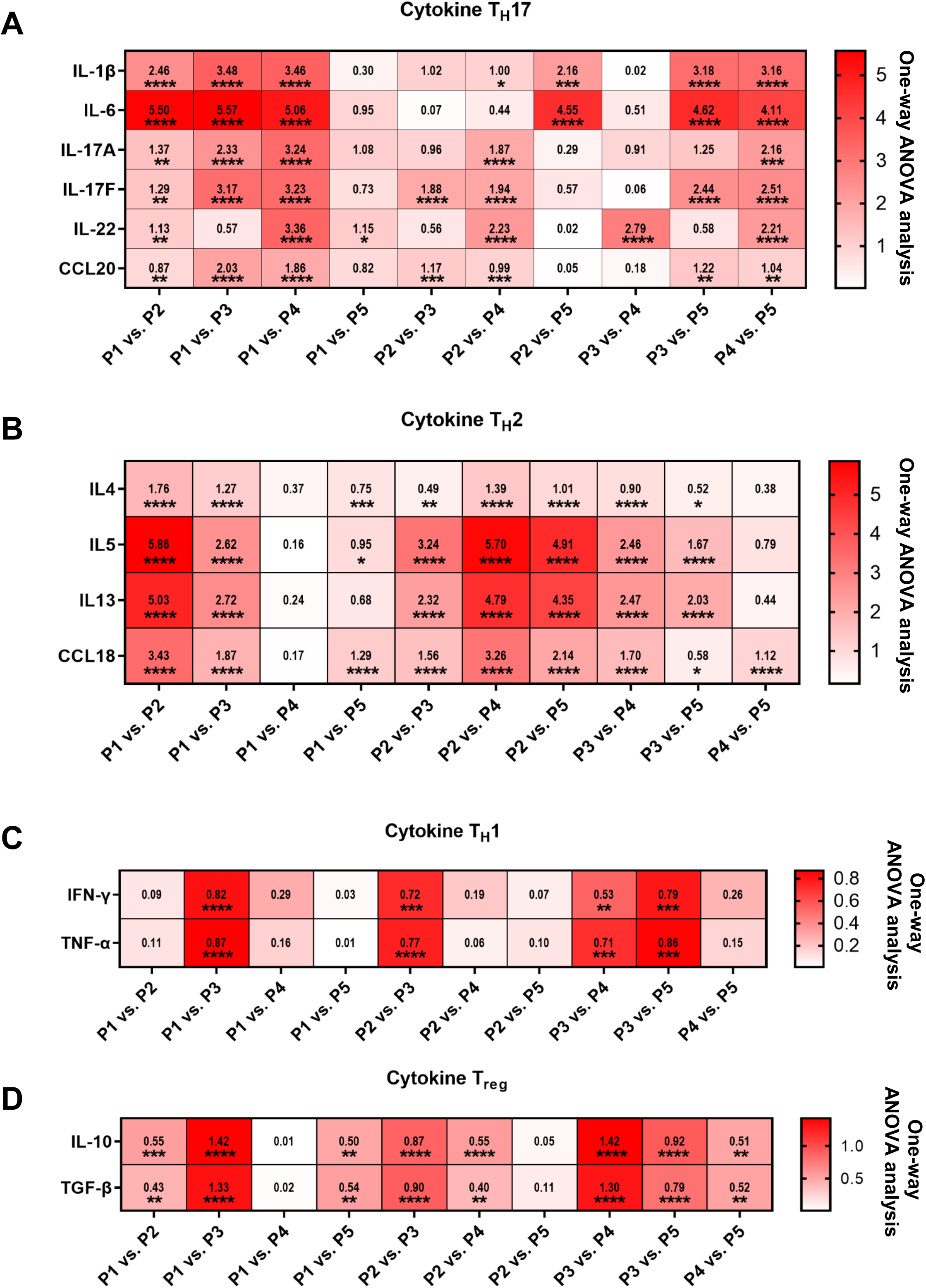
Two-way Anova analysis of cytokine expression levels across the 5 CI dysbiosis clusters. Heatmap diagrams showing relative expression of Th17 (A), Th2 (B), Th1 (C), and Treg (D) cytokine levels for each CI group (P2-P5) compared to healthy controls (P1), and each CI group compared each other. *\*p* ≤ 0.05, *\*\*p* ≤ 0.01, *\*\*\*p* ≤ 0.001, *\*\*\*\*p* ≤ 0.0001

**Figure S9.**
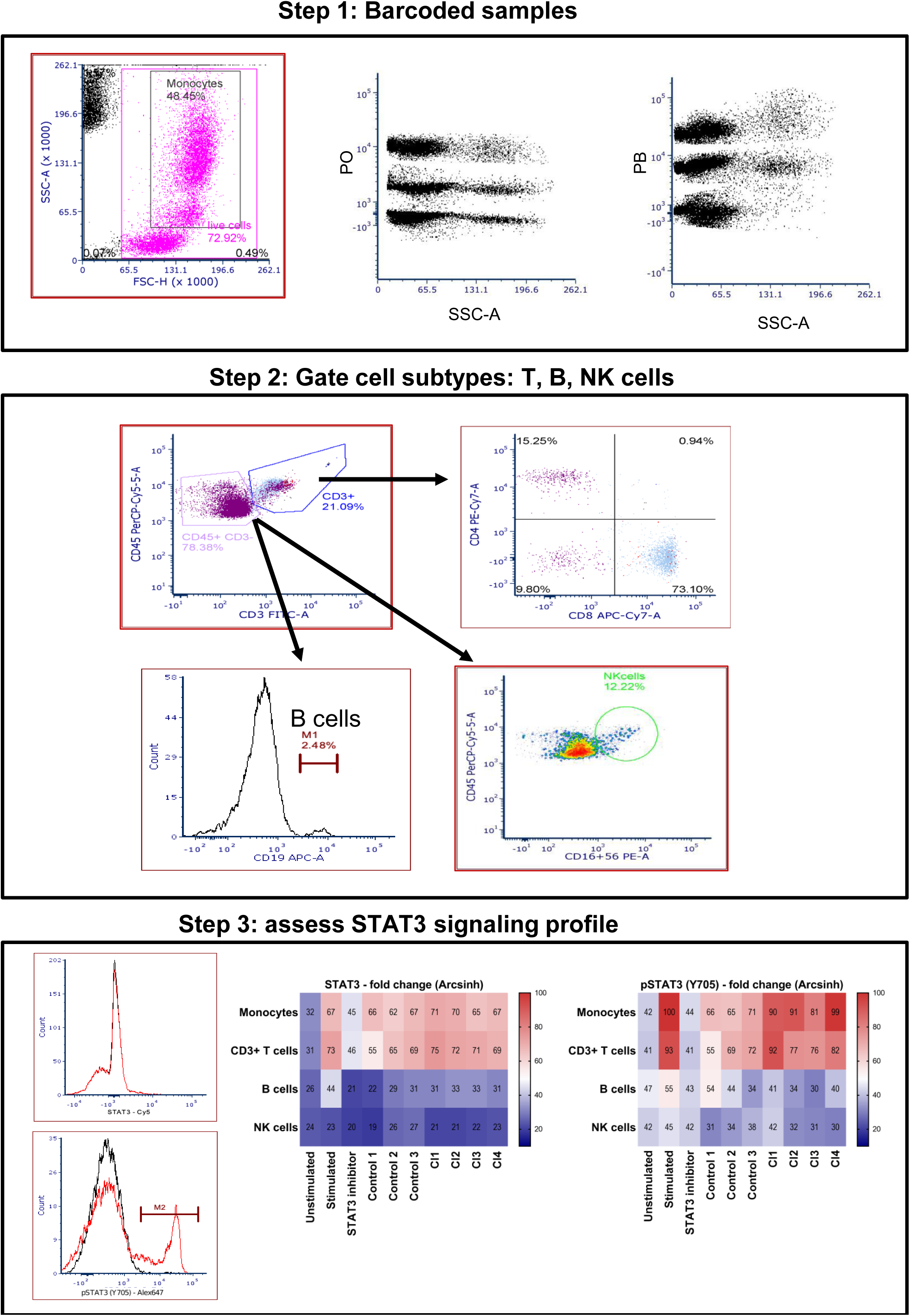
Procedure for determining JAK/STAT signaling profile in CI patients. High-throughput subtyping of cells via flow cytometry enabled assessment following the protocol of Davies et al., 2016. Cryopreserved PBMCs were thawed and rested at 37 °C with serum free media, 5% CO2 for 11.5 h to reduce basal phosphorylation levels before plated into Megablock® 96 well plate in a final cell concentration of 1 × 106 cells/ml. Control samples were unstimulated or stimulated for 15 min with 400 μl Combo (cytokines (100 ng/ml), LPS (10 μg/ml; Sigma-Aldrich) or PMA (100 ng/ml; Sigma-Aldrich)). The patient samples were unstimulated by any substances. Next, samples were fixed, permeabilized, and barcoded with staining of pacific blue (PB) and pacific orange (PO) dyes. Single cells were gated based on their forward scatter area (FSC-A) and forward scatter width (FSC-W), followed by intact cells based on side scatter area (SSC-A) and FSC-A. The different stimulation conditions were then identified through the intensities of their PB and PO stains. Cell subtypes were identified based on their FSC-A and SSC-A scatter properties as either monocytes or lymphocytes. Lymphocytes were then subtyped based on surface antigens including B cells (CD19 +), T cells (CD3 +/ CD4 +, CD8 + CD56 −), NK cells (CD3 − CD56 +). Cells within each subtype were analysed based on the change of normalized median fluorescence intensity in each stimulation condition relative to the unstimulated reference samples. STAT3 and STAT3 pY705 levels in monocytes, CD3+ T-cells, B cells, and NK cells were analysed in scatter graphs and heat maps.

**Figure S10.**
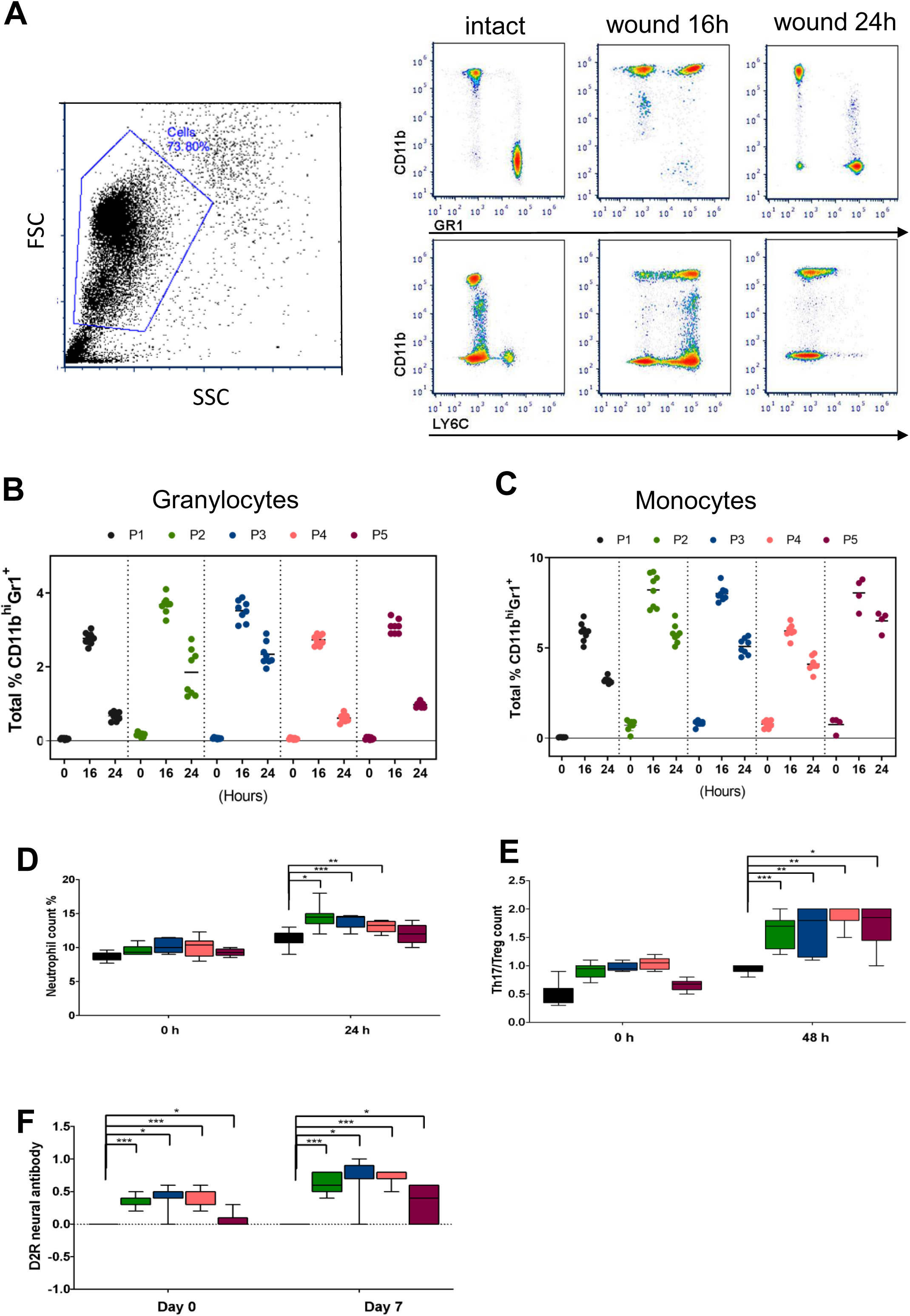
Relative granulocyte and monocyte inflammation in CI patients following wounding of skin. A. Flow cytometry gating strategy for CD11b^hi^, GrR1^+^ granulocytes, and CD11b^hi^, Ly6C^+^ monocytes B. Relative CD11b^hi^Gr1^+^ granulocyte populations followed longitudinally over 24 hours following wounding for all 5 CI dysbiosis clusters. Results expressed in dot plot with each horizontal bar representing the mean of the group. C. Relative CD11b^hi^Ly6C^+^ monocyte populations followed longitudinally over 24 hours following wounding for all 5 CI dysbiosis clusters. Results expressed in dot plot with each horizontal bar representing the mean of the group. D. Percentage of neutrophils counted over 24 hours compared in each CI dysbiosis population following wounding. Results are expressed in dot plot showing median and interquartile range with statistical significance as *\*p* ≤ 0.05, *\*\*p* ≤ 0.01, *\*\*\*p* ≤ 0.001, *\*\*\*\*p* ≤ 0.0001. E. Percentage of Th17/Treg cells counted over 48 hours compared in each CI dysbiosis population following wounding. Results are expressed in dot plot showing median and interquartile range with statistical significance as *\*p* ≤ 0.05, *\*\*p* ≤ 0.01, *\*\*\*p* ≤ 0.001, *\*\*\*\*p* ≤ 0.0001. F. Number of auto-antibody D2R counted over 7 days compared in each CI dysbiosis population following wounding. Results are expressed in dot plot showing median and interquartile range with statistical significance as *\*p* ≤ 0.05, *\*\*p* ≤ 0.01, *\*\*\*p* ≤ 0.001, *\*\*\*\*p* ≤ 0.0001.

