## Supplemental Table 2 for "Altered skin microbiome, inflammation, and JAK/STAT signaling in Southeast Asian ichthyosis patients"

**Supplemental table 2: result of microbial realtime PCR**

### A) Bacterial infection assay Realtime PCR

Nasopharyngeal swab

| cohort 1 | S.pneumoniae | M.Catarrhalis | H.influenzae | S.pyogenes | S.agalacitae |
|---|---|---|---|---|---|
| HI1 | 29 | 28.6 | 33 | 40 | 39.1 |
| HI5 | 33.2 | 32.1 | 35 | 30.4 | 32.9 |
| HI8 | 32.7 | 30.09 | 32.5 | 33.2 | 35.4 |
| IV1 | 28.5 | 33.2 | 35.4 | 35.1 | 33.7 |
| IV2 | 22.4 | 32.6 | 32.6 | 35.7 | 40.1 |
| IV3 | 32.7 | 32.6 | 35.4 | 40.1 | 39.5 |
| IV7 | 33.8 | 33.2 | 30.6 | 33.2 | 39.6 |
| IV9 | 33.9 | 32.5 | 30.3 | 33.4 | 29.4 |
| IV10 | 34.6 | 30.6 | 30.5 | 30.9 | 30.1 |
| IV11 | 35.4 | 32.6 | 34.7 | 33.4 | 39.7 |
| IV12 | 37.5 | 33.5 | 33.2 | 33.5 | 39.2 |
| IV13 | 31.6 | 32.4 | 38.7 | 30.6 | 39.3 |
| IV14 | 33.2 | 25.4 | 39.5 | 34.7 | 39.1 |
| IV15 | 29.8 | 35.7 | 35.6 | 33.2 | 33.2 |
| EI1 | 29.6 | 30.7 | 36.8 | 38.7 | 33.5 |
| EI2 | 30.7 | 34.7 | 39 | 39.5 | 33.1 |
| TTD1 | 29.8 | 39.5 | 30.9 | 38.7 | 40 |
| TTD2 | 33.4 | 28.5 | 40.2 | 40 | 39.4 |
| TTD3 | 33.2 | 29.4 | 40.1 | 41.2 | 37.9 |
| SLS1 | 31.8 | 32.5 | 40 | 33.6 | 37.6 |
| Threshold positive | ≥ 33 | ≥ 34 | ≥ 40 | ≥ 40 | ≥ 40 |

**postive**

**below basal/negative**

### B) Antibiotic assay realtime PCR

Nasopharyngeal swab

| cohort 1 | Sulfonamide | Trimethoprim | Beta-lactam | Tetracycline |
|---|---|---|---|---|
| HI1 | 1 | 0 | 1 | 0 |
| HI5 | 0 | 1 | 0 | 0 |
| HI8 | 0 | 0 | 0 | 0 |
| IV1 | 0 | 0 | 0 | 0 |
| IV2 | 0 | 0 | 1 | 0 |
| IV3 | 1 | 0 | 0 | 0 |
| IV7 | 0 | 0 | 0 | 0 |
| IV9 | 0 | 0 | 1 | 0 |
| IV10 | 0 | 0 | 1 | 0 |
| IV11 | 0 | 0 | 0 | 0 |
| IV12 | 0 | 0 | 0 | 0 |
| IV13 | 0 | 0 | 0 | 0 |
| IV14 | 0 | 0 | 0 | 0 |
| IV15 | 0 | 0 | 0 | 1 |
| EI1 | 0 | 0 | 0 | 0 |
| EI2 | 0 | 0 | 1 | 0 |
| TTD1 | 0 | 0 | 0 | 0 |
| TTD2 | 0 | 0 | 1 | 0 |
| TTD3 | 0 | 0 | 0 | 0 |
| SLS1 | 0 | 0 | 0 | 0 |
| Threshold positive | 1 | 1 | 1 | 1 |

Skin specimen

| cohort 1 | S.aureus |
|---|---|
| HI1 | 25 |
| HI5 | 28 |
| HI8 | 29.3 |
| IV1 | 28.5 |
| IV2 | 28.2 |
| IV3 | 35.5 |
| IV7 | 29.3 |
| IV9 | 38.7 |
| IV10 | 39.2 |
| IV11 | 29.4 |
| IV12 | 31.5 |
| IV13 | 33.1 |
| IV14 | 28.2 |
| IV15 | 28.4 |
| EI1 | 33.1 |
| EI2 | 27.5 |
| TTD1 | 26.3 |
| TTD2 | 29.4 |
| TTD3 | 29.1 |
| SLS1 | 28.2 |
| Threshold posit | ≥ 28 |

### C) Viral detection assay realtime PCR

Skin, gental specimen

| cohort 2 | HPV general | HPV16 | HPV18 | HPV6 | Herpesvirus | MCV |
|---|---|---|---|---|---|---|
| HI1 | 0 | 0 | 0 | 0 | 0 | 0 |
| HI2 | 0 | 0 | 0 | 0 | 0 | 0 |
| HI3 | 0 | 0 | 0 | 0 | 0 | 0 |
| HI4 | 0 | 0 | 0 | 0 | 0 | 0 |
| HI5 | 1 | 0 | 0 | 0 | 1 | 0 |
| HI6 | 0 | 0 | 0 | 0 | 0 | 0 |
| HI7 | 0 | 0 | 0 | 0 | 1 | 0 |
| HI8 | 0 | 0 | 0 | 0 | 0 | 0 |
| LI1 | 0 | 0 | 0 | 0 | 0 | 0 |
| LI2 | 1 | 0 | 0 | 0 | 0 | 0 |
| LI3 | 0 | 0 | 0 | 0 | 1 | 0 |
| IV1 | 0 | 0 | 0 | 0 | 1 | 0 |
| IV2 | 0 | 0 | 0 | 0 | 1 | 0 |
| IV3 | 1 | 0 | 0 | 0 | 0 | 0 |
| IV4 | 1 | 1 | 0 | 0 | 0 | 0 |
| IV5 | 0 | 0 | 0 | 0 | 1 | 0 |
| IV6 | 0 | 0 | 0 | 0 | 0 | 0 |
| IV7 | 1 | 0 | 0 | 0 | 1 | 0 |
| IV8 | 1 | 0 | 0 | 0 | 1 | 0 |
| IV9 | 1 | 1 | 0 | 0 | 0 | 1 |
| IV10 | 1 | 1 | 0 | 0 | 1 | 0 |
| IV11 | 1 | 0 | 0 | 0 | 1 | 0 |
| IV12 | 0 | 0 | 0 | 0 | 0 | 0 |
| IV13 | 0 | 0 | 0 | 0 | 1 | 0 |
| IV14 | 1 | 0 | 0 | 1 | 0 | 1 |
| IV15 | 0 | 0 | 0 | 0 | 1 | 0 |
| EI1 | 0 | 0 | 0 | 0 | 1 | 0 |
| EI2 | 0 | 0 | 0 | 0 | 0 | 0 |
| EI3 | 1 | 0 | 0 | 0 | 1 | 0 |
| EI4 | 1 | 0 | 0 | 0 | 1 | 0 |
| TTD1 | 1 | 0 | 0 | 0 | 1 | 0 |
| TTD2 | 1 | 1 | 0 | 0 | 1 | 0 |
| TTD3 | 1 | 1 | 0 | 0 | 1 | 0 |
| ARC1 | 0 | 0 | 0 | 0 | 1 | 0 |
| SLS1 | 0 | 0 | 0 | 0 | 0 | 0 |
| SLS2 | 1 | 0 | 0 | 0 | 1 | 0 |
| HT1, relative of HI | 0 | 0 | 0 | 0 | 0 | 0 |
| HT2, relative of IV | 0 | 0 | 0 | 0 | 0 | 0 |
| HT3, relative of TTD | 0 | 0 | 0 | 0 | 0 | 0 |
| HT4, relative of LI | 0 | 0 | 0 | 0 | 0 | 0 |
| HT5, relative of IV | 0 | 0 | 0 | 0 | 0 | 0 |
| HT6, relative of ARC | 1 | 0 | 0 | 0 | 1 | 0 |
| HT7, volunteer | 1 | 0 | 0 | 0 | 1 | 0 |
| | 22 | 7 | 0 | 1 | 27 | 2 |
| PATIENT | 42.55319149 | 14.89361702 | 0 | 2.127659574 | 51.06382979 | 4.255319149 |
| HC | 28.57142857 | 0 | 0 | 0 | 28.57142857 | 0 |
