## Supplemental Table 3 for "Altered skin microbiome, inflammation, and JAK/STAT signaling in Southeast Asian ichthyosis patients"

**Supplement table 3: Clinical patterns of heat loss, erythroderma**

| Patient ID | Erythroderma in total body | Erythroderma in local lesion | Cutaneous heat loss (<30-32) | Onset of age diagnosis |
|---|---|---|---|---|
| HI1 | yes | yes | NA | 0-5 years |
| HI2 | yes | yes | NA | 0-5 years |
| HI3 | yes | yes | yes | 0-5 years |
| HI4 | yes | yes | NA | 0-5 years |
| HI5 | yes | yes | yes | 0-5 years |
| HI6 | yes | yes | yes | 0-5 years |
| HI7 | yes | yes | NA | 0-5 years |
| HI8 | yes | yes | NA | 0-5 years |
| LI1 | no | yes | YES | 11-15 years |
| LI2 | no | no | NA | 0-5 years |
| LI3 | no | no | NA | 0-5 years |
| IV1 | no | yes | NA | 0-5 years |
| IV2 | no | yes | no | 6-10 years |
| IV3 | no | no | no | 16-20 years |
| IV4 | no | no | no | 11-15 years |
| IV5 | no | yes | no | 6-10 years |
| IV6 | no | yes | no | 6-10 years |
| IV7 | no | yes | no | 16-20 years |
| IV8 | no | yes | no | 16-20 years |
| IV9 | no | no | no | 61-65 years |
| IV10 | no | yes | no | 16-20 years |
| IV11 | no | no | no | 26-30 years |
| IV12 | no | yes | no | 0-5 years |
| IV13 | no | yes | no | 0-5 years |
| IV14 | no | no | no | 41-45 years |
| IV15 | no | yes | no | 0-5 years |
| EI1 | no | yes | NA | 0-5 years |
| EI2 | no | yes | yes | 16-20 years |
| EI3 | no | yes | yes | 6-10 years |
| EI4 | no | yes | no | 21-25 years |
| WN1 | no | yes | NA | 16-20 years |
| WN2 | no | yes | NA | 0-5 years |
| WN3 | no | yes | NA | 0-5 years |
| DI1 | no | no | no | 11-15 years |
| DI2 | no | no | NA | 0-5 years |
| DKH1 | no | no | no | 41-45 years |
| DKH2 | no | no | no | 6-10 years |
| DKH3 | no | no | no | 31-35 years |
| DKH4 | no | no | no | 6-10 years |
| DKH5 | no | no | no | 6-10 years |
| DKH6 | no | no | no | 6-10 years |
| TTD1 | no | no | no | 36-40 years |
| TTD2 | no | yes | no | 26-30 years |
| TTD3 | no | yes | no | 26-30 years |
| ARC1 | no | no | no | 0-5 years |
| SLS1 | no | yes | no | 6-10 years |
| SLS2 | no | no | no | 6-10 years |

*Loss of heat: the measurement was examined by the infrared Thermal Imaging Camera (2.4 Inch 24*32 Resolution) Instrument (VIM-640G2ULC, Vision Sensing, Japan)
